# A multi-layered approach to elucidate mechanisms of physical function in response to rehabilitation in heart failure with preserved ejection fraction

**DOI:** 10.64898/2026.02.12.26346203

**Authors:** Andrew S. Perry, Christopher O’Connor, Mirko Pavicic, Quanhu Sheng, Eric Farber-Eger, Aditya Sarkar, Phillip Lin, Patrick Evans, Kahraman Tanriverdi, Antonina Risitano, Anthony E. Peters, Haiying Chen, Bharathi Upadhya, David J. Whellan, Amy Pastva, Robert J. Mentz, Alain Bertoni, Charles Semelka, Peter H Brubaker, Anthony J A Molina, Russ Newland, Ben Nelson, Matthew Lane, Kyle A. Sullivan, Alice Townsend, Anna Vlot, Bryan Nsoh, Paul D. Allen, Quinn Wells, Gordon R. Reeves, Daniel Jacobson, Dalane W. Kitzman, Eric R. Gamazon, Matthew Nayor, Ravi V. Shah

## Abstract

Heart failure with preserved ejection fraction (HFpEF) is an increasingly common cause of morbidity and mortality in older adults that is driven by cardiac and non-cardiac mechanisms. Physical rehabilitation improves frailty and functional capacity in HFpEF, though underlying mechanisms remain less clear. We quantified >5,000 circulating proteins across two randomized clinical trials of rehabilitation in HFpEF (REHAB-HF, SECRET-II), identifying proteins associated with prognostic measures of physical function (short physical performance battery, 6-minute walk distance) and protein changes after rehabilitation. Using an artificial intelligence (AI)-enabled multiplex network analysis (MENTOR-IA), we identified biologically plausible networks central to this “physical function proteome,” including endothelial remodeling, mitochondrial metabolism, calcium handling, and immune modulation. Expression of prioritized proteins at the transcriptional level localized to heart, skeletal muscle, and brain tissue, with several cognate transcripts implicated in frailty via tissue-specific transcriptome-wide genetic association studies. In addition, using novel human genetic approaches, we implicated select proteins as mediating tissue-specific genetic effects on frailty. These findings motivated us to construct multi-protein signatures of physical function, which correlated with functional changes observed with rehabilitation in REHAB-HF and SECRET-II and that were associated with heart failure and multi-dimensional clinical outcomes in >26,000 individuals. These findings collectively delineate a multi-system molecular program underlying physical function impairment and rehabilitation response in HFpEF, offering insights into potential precision risk estimators and therapeutic targets for surveillance and promotion of physiologic resilience.

## INTRODUCTION

Heart failure (HF) with preserved ejection fraction (HFpEF) is the most common form of heart failure among older persons, with impaired physical function as a cardinal consequence driving poorer quality of life^1,2^. Unfortunately, numerous pharmacological targets have been evaluated across several large, multi-center randomized controlled trials which have largely yielded negative results. In contrast, multiple randomized controlled trials have consistently demonstrated that exercise training and physical rehabilitation improve physical function and quality of life in HFpEF^3^, even when initiated early during the index hospitalization for HF decompensation. Although exercise and rehabilitation consistently provide the largest improvement in physical function and quality of life across different treatments in HFpEF (including pharmacological), *how* rehabilitation improves function in HFpEF—critical to targeting at-risk populations with HFpEF and finding modifiable pathways to improve function*—*is less clear. In addition, unique challenges to studying rehabilitation in older adults with HFpEF—including availability of large studies of rehabilitation or access to tissue—have limited our ability to interrogate underlying molecular mechanisms responsible for the corresponding functional improvements. Understanding mechanisms linked to clinically meaningful changes in physical function in HFpEF could inform enhanced rehabilitation strategies, novel targeted pharmacological intervention, and clinically useful biomarkers to stratify risk in this vulnerable population.

Here, we quantified a circulating proteome in 214 individuals across two recently completed randomized controlled trials of physical intervention in acute decompensated HFpEF (REHAB-HF; early, transitional, tailored, structured, and progressive multidomain physical rehabilitation intervention) and chronic stable HFpEF (SECRET-II; resistance training added to aerobic training and caloric restriction). We hypothesized that a “physical function proteome”—defined as proteins linked to prognostic physical function phenotypes targeted by rehabilitation (short physical performance battery, 6-minute walk distance) and those that change during rehabilitation intervention—would inform physiology of functional reserve in HFpEF, tissue-specific mechanisms of frailty, and long-term clinical morbidity. Our approach leveraged these two randomized controlled studies of rehabilitation in HFpEF to identify proteins linked to functional traits or that are dynamic with intervention (physical function proteome). We subsequently characterized the physiologic relevance of this physical function proteome via standard and novel agentic artificial intelligence (AI)-based approaches, bulk and single cell transcriptomics in target tissues of interest, and novel human genetic inferential methods. The mechanistically plausible impact of the physical function proteome on HFpEF biology from these approaches prompted us to examine its relations with 22 multi-system outcomes frequently encountered in older adults with HFpEF in >26,000 individuals, demonstrating its prognostic capability in HFpEF and related comorbidity. Ultimately, by integrating randomized clinical trials in HFpEF with transcriptomics, high-performance AI supercomputing, new tissue-specific human genetics and population-based studies, we present a comprehensive clinical-biological study of physical rehabilitaiton in HFpEF, pinpointing putative mechanisms of physical function and biomarkers that mark liability to clinical outcome.

## METHODS

### Overview structure of the study

The overall flow of the study is displayed in **Figure 1**. As a first step, we defined a “physical function proteome” by identifying proteins in circulation linked to two functional endpoints—short physical performance battery (SPPB) and 6-minute walk distance (6MW)—or change with rehabilitation intervention in two randomized controlled trials (REHAB-HF, SECRET-II). For SPPB- and 6MW-associated proteins, we replicated protein-phenotype associations against frailty phenotypes in a separate study of age-related frailty in severe aortic stenosis^4^ and in a large population-based study with measures of frailty (UK Biobank). To clarify biological relevance of the physical function proteome (our second goal), we next used standard bioinformatics methods and novel agentic AI-based supercomputing to identify linked biological pathways. We queried available bulk and single cell transcriptomic reference data (including data across age and sarcopenia status) to understand transcriptional origin of the implicated proteome and its dynamicity with HFpEF-relevant states. We also deployed established and developed new genomic methods to test how transcripts encoding the physical function proteome may encode tissue-specific genetic liability to frailty, a major condition limiting physical function in our randomized controlled trials. These studies prompted our third goal, to understand the clinical relevance of a physical function-informed proteome on HFpEF-relevant morbidity. Given the functional limitations in HFpEF are multi-organ, we constructed multi-protein signatures of SPPB and 6MW and tested their ability to predict response to rehabilitation intervention and their relation to 22 clinical outcomes in >26,000 individuals in the population (UK Biobank).

**Figure 1.**
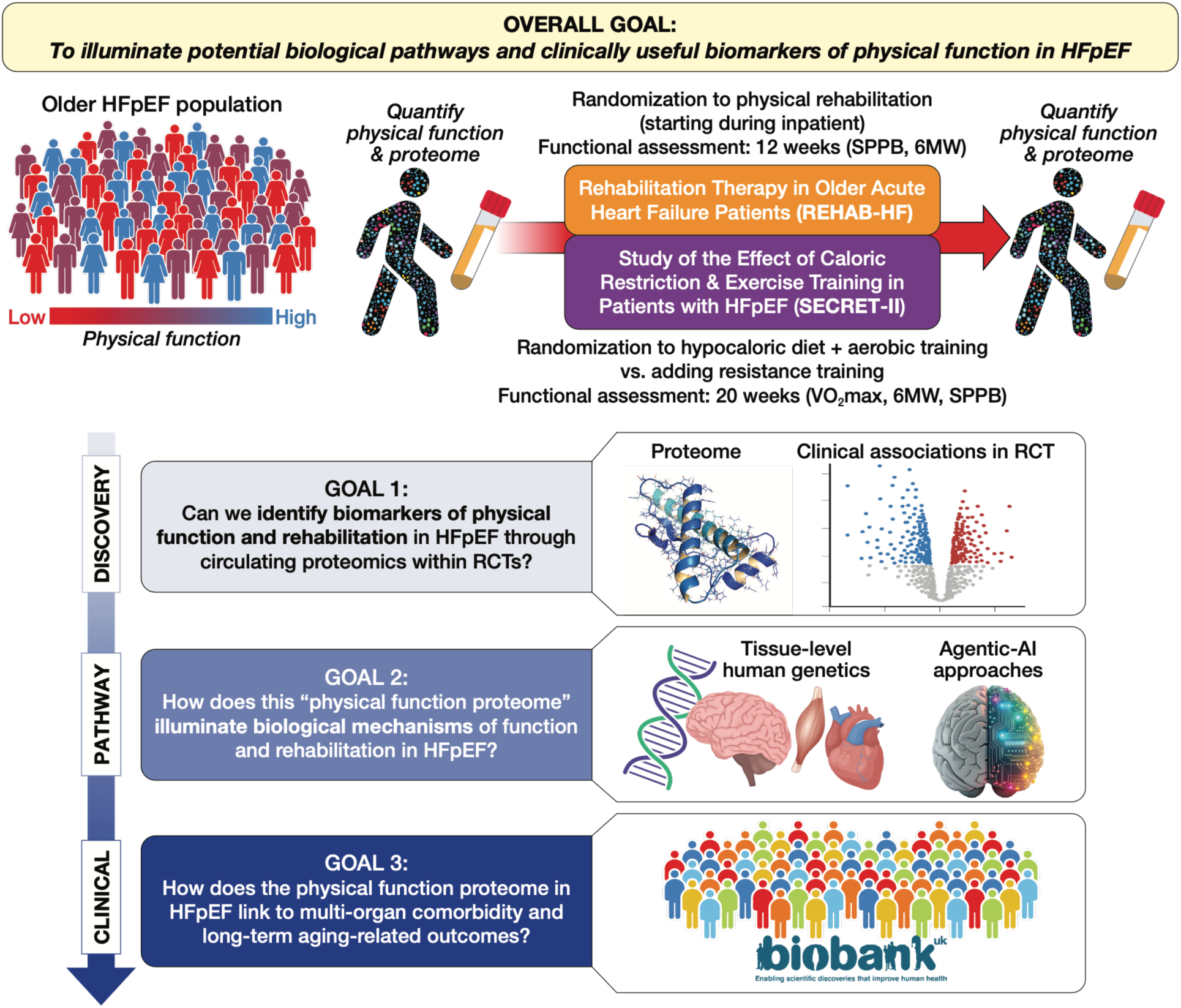
Graphical abstract.

### Data availability

Data from the REHAB-HF and SECRET-II trials are available on request from the study coordinating centers (via contact to author DWK). Data from the UK Biobank are available through the research analysis platform (https://www.ukbiobank.ac.uk/use-our-data/research-analysis-platform/). This research has been conducted using the UK Biobank Resource under Application Number 57492. Additional data (aortic stenosis cohort, RNA-seq data, GWAS) are available through the references^4–8^.

### Code Availability

Analytical code for this study is available at https://github.com/asperry125/HFpEF-Rehabilitation-Proteomics.git.

### Study cohorts

We leveraged data from two randomized controlled trials (REHAB-HF, SECRET-II) and a population-based cohort (UK Biobank). Details of the randomized trials procedures and description have been reported previously with key aspects repeated here (attribution to prior reporting given by this statement)^9–12^.

#### Rehabilitation Therapy in Older Acute Heart Failure Patients (REHAB-HF)

The REHAB-HF randomized controlled trial tested whether an early, transitional, tailored, structured, and progressive multidomain physical rehabilitation intervention initiated early during hospitalization, when feasible, and continued after discharge through outpatient and home-based phases would improve physical function and reduce all-cause rehospitalization in older patients hospitalized for acute decompensated heart failure. Details have been reported previously with key aspects repeated here (attribution to prior reporting given by this statement)^9–11^. In brief, patients hospitalized for HF were eligible if they were ≥60 years old, able to complete the physical assessment, and anticipated to be discharged home post-hospitalization. Participants were randomized to either (1) rehabilitation intervention or (2) usual care. Participants randomized to physical rehabilitation began the intervention prior to hospital discharge when feasible and continued with outpatient sessions of 1 hour, 3 days/week, for 12 weeks (36 total sessions). Participants randomized to usual care received phone calls every 2 weeks in addition to usual care. All participants had in-person visits at 1 and 3 months. A pre-planned analysis by LVEF subgroups found that participants with HFpEF (defined as left ventricular ejection fraction [LVEF] ≥45%), received disproportionately greater benefit from the rehabilitation intervention and are included in this study (**Supplemental Table 1**)^13^. Patients who did not have any available blood samples for proteomics were excluded (blood collection not mandated in REHAB-HF).

#### Study of the Effects Caloric Restriction and Exercise Training in Patients With Heart Failure and a Normal Ejection Fraction (SECRET-II)

The SECRET-II randomized controlled trial tested whether addition of resistance training to a combination of aerobic training and caloric restriction would further improve physical function by mitigating loss of skeletal muscle observed with caloric restriction^14^. Details have been published previously with key aspects repeated here (attribution of prior reporting given by this statement^12^). Participants were eligible for SECRET-II if they had HFpEF (LVEF≥50%), were age ≥60 years, and had a body mass index (BMI) ≥28kg/m^2^. All participants were prescribed a low-calorie diet (meals prepared by the Wake Forest Clinical Research Unit Metabolic Kitchen) and aerobic exercise training (40 minutes, 3 times/week) for 20 weeks. Participants randomized to resistance training added 20 minutes of resistance exercise, while other participants added an additional 20 minutes of recovery exercise to this regimen. Participants underwent symptom limited cardiopulmonary exercise testing (CPET; Modified Naughton Protocol).

#### UK Biobank

UK Biobank (UKB) is a population-based epidemiologic study encompassing ≈500,000 individuals age 40-69 years at recruitment (2006-2010). We included UKB participants with circulating proteomics quantification as part of the UK Biobank Pharma Proteomics Project (≈52,000 participants)^15^.

### Physical function assessment

The primary measures of physical function were the SPPB and 6MW. Both of these standardized measures are clinically meaningful, and are associated with frailty, QOL and clinical events^16,17^. SPPB is comprised of 3 tests: (1) standing balance; (2) gait speed quantification (4-m walk); (3) strength assessment (time needed to rise from a chair 5 times)^18^. Each test is graded on a 0-4 scale, with lower sum scores (range 0-12) indicating worse physical function. SPPB and 6MW were assessed in both trials using similar protocols. Physical function in UKB was assessed via questionnaire and handgrip strength^19^. We calculated the Fried frailty score in UKB using previously reported methods^20^. We used questionnaire responses to determine weight loss over the preceding year (UKB Data Field 2306), frequency of tiredness in the preceding 2 weeks (exhaustion; UKB Data Field 2080), walking speed (UKB Data Field 924), average handgrip strength (weakness; UKB Data Fields 46, 47), and physical activity (UKB Data Fields 6164, 1011).

### Proteomics

We employed the Olink Explore HT platform (5,416 unique proteins) in REHAB-HF and SECRET-II. Methods for Olink Explore-based protein quantification have been previously reported^21^. We quantified proteomics in all included samples in REHAB-HF and SECRET-II in the same batch to minimize technical variability. We filtered our initial data by (1) excluding participants with >500 missing protein measurements (N=8); (2) excluding participants identified as outliers by principal component analysis (N=1, outlier defined as being >5 standard deviations away from the mean); (3) excluding proteins with any missing measurements (N=202). Transformations to proteomics are described as relevant throughout the **Statistical analysis**. The final analytic sample included 214 participants with 5,214 unique circulating proteins. Proteomics data from UK Biobank was generated by the Pharma Proteomics Project which used the Olink Explore 3072 platform^15^.

### Statistical analysis

A key aspect of our analyses is a limited sample size to number of proteins in the REHAB-HF and SECRET-II randomized controlled trials. Our approach therefore relied on merging SECRET-II and REHAB-HF (for cross-sectional, pre-intervention analyses) and considering trials separately (for longitudinal analyses). To balance the potential for discovery within our clinical trials with the need to control type 1 error, we deployed more lenient type 1 error thresholds in REHAB-HF and SECRET-II and a more stringent type 1 error in subsequent replication proteomic and human genetic analyses. In addition, we used machine learning methods specifically designed for the imbalance between sample size and predictor space through penalized regression modeling (as in our previous work^4^). Finally, we replicated associations in other cohorts of HF and multi-morbidity to enhance generalizability^4,22^. The overall flow of our analyses (including the thresholds used for discovery and replication) is shown in **Supplemental Figure 1**.

#### Identifying proteins associated with pre-intervention SPPB and 6MW

We first estimated age-/sex-adjusted linear models for each outcome (SPPB, 6MW) as a function of each protein individually (type 1 error controlled by Benjamini-Hochberg false discovery rate [FDR]<10%). Given we were concerned with pre-intervention phenotypes in this step, we merged REHAB-HF and SECRET-II in these models. For these models, protein data were normalized (mean centering, unit variance) using the mean and variance of the merged dataset. To examine replication and generalizability, we compared protein effect sizes in two additional cohorts with ascertainment of functional outcomes: (1) advanced CVD—809 individuals with severe aortic stenosis where grip strength and gait speed were assessed (with Olink Explore 1536 proteomics)^4^ and (2) community-dwelling individuals—UKB participants with available measures of handgrip strength and Fried frailty score. Targets identified here were carried into downstream analyses (see “Linking dynamic proteomics of physical function to aging-related multi-morbidity,” “Standard and artificial intelligence (AI)-enabled network and pathway interrogation,” “Localizing targets in the physical function proteome at tissue- and single cell-level resolution through transcriptomics,” and “Genomic approaches to tissue-specific liability to frailty and proteogenomic integration”).

#### Defining and characterizing a multi-protein signature of SPPB and 6MW

To accommodate the sample size-to-protein discrepancy and generate a *clinically translatable* signature of functional impairment in HFpEF, we used least absolute shrinkage and selection operator regression (LASSO) to define multivariable proteomic signatures of <u>pre</u>-intervention SPPB and 6MW, including all 5,214 proteins. We used cross-validation for hyperparameter optimization^23^. We merged REHAB-HF and SECRET-II for these analyses to maximize power across a broad range of SPPB and 6MW encompassed by REHAB-HF and SECRET-II participants (**Supplementary Figure 2**). For LASSO models, protein data were normalized (mean centered, unit variance) using the mean and variance calculated from a merged dataset including both trial cohorts and both timepoints (baseline, 3-month follow up). Fit of the LASSO-defined multi-protein signatures against SPPB and 6MW was tested in each trial separately. In addition, we examined changes in this multi-protein signature during trial intervention and tested the fit of models built on the pre-intervention phenotypes against <u>post</u>-intervention SPPB and 6MW (using post-intervention protein levels). Finally, we used analysis of variance (type 1 ANOVA) to quantify contribution of key clinical determinants to each multi-protein signature (age, sex, BMI, hypertension, atrial fibrillation, diabetes, smoking, and estimated glomerular filtration rate).

#### Examining changes in the proteome with intervention

We next assessed changes in the proteome with intervention *separately* in REHAB-HF and SECRET-II (given differences in the type of intervention in each trial). We specified linear models as *follow-up protein level ∼ baseline protein level + randomization status + age + sex*, with model coefficient on <u>randomization</u> term interpreted as an estimated mean effect of randomized group on follow-up protein level (intervention vs. control arm). Protein data were not transformed for these models to allow for the model coefficients to be interpreted as an adjusted log_2_ fold change. To optimize discovery potential, we used a nominal 5% significance level to nominate targets for downstream evaluation (see “Linking dynamic proteomics of physical function to aging-related multi-morbidity,” “Standard and artificial intelligence (AI)-enabled network and pathway interrogation,” “Localizing targets in the physical function proteome at tissue- and single cell-level resolution through transcriptomics,” and “Genomic approaches to tissue-specific liability to frailty and proteogenomic integration”).

#### Linking dynamic proteomics of physical function to aging-related multi-morbidity

We evaluated association of (1) multi-protein signatures of SPPB and 6MW and (2) individual proteins *dynamic* during intervention from REHAB-HF and SECRET-II with age-relevant multi-morbidity in 26,263 UKB participants. Methods described here are analogous to our prior methods (attribution of prior reporting by this statement^24^). Given differences in protein coverage between the two trials and UKB (Olink Explore HT vs. Olink Explore 3072), we recalibrated models from the trials via LASSO regression in REHAB-HF/SECRET-II, akin to prior approaches^24^. In this approach, we first predicted SPPB and 6MW using 5,214 proteins in REHAB-HF and SECRET-II based on our initial LASSO model. We then specified a LASSO model for these protein-predicted values as the outcome and 2,645 proteins found in both UKB and REHAB-HF/SECRET-II (matched by assay name) as predictors for selection. This approach generates model coefficients for a signature that can be applied to UKB for regression built on the initial predictions in REHAB-HF and SECRET-II. We compared the initial “full” model predictions (across 5,214 proteins) with the “recalibrated” model (across the 2,645 overlap proteins) via Spearman correlation coefficient. We moved these “recalibrated” signatures and individual proteins that changed with intervention (at nominal P<5% significance level, matched by assay with UKB) forward.

Given the multi-organ origin of functional limitation and morbidity in HFpEF, we constructed Cox models in UKB for key age- and multi-organ outcomes, adjusted for age, sex, race/ethnicity, BMI, hemoglobin A1c, triglycerides, high-density lipoprotein (HDL), smoking, alcohol use, and Townsend Deprivation Index. Protein signatures (from the recalibrated models) and individual proteins were used as predictors. Outcomes were defined by the PheWAS package^25^. Prevalent cases were excluded from models; prevalent cases were defined by self-report questionnaires of diseases or related conditions (UKB Data Fields 2443, 20002), serum chemistry data where applicable (e.g., A1c thresholds for diabetes), and presence of a pre-existing or confounding ICD code at the time of recruitment. Time-to-death data was defined by registry data (UKB Data Field 40000), and non-deaths were censored on 30 November 2022. Time-to-event data for non-death outcomes was defined as the time to first qualifying ICD code, and non-event participants were censored at the date of death (if applicable) or region-specific dates (UKB Data Field 54): 31 October 2022 for England; 31 July 2021 for Scotland; and 28 February 2018 for Wales. In addition to Cox models, we also quantified the correlation of the recalibrated signatures against handgrip and Fried frailty score in UKB.

### Standard and artificial intelligence (AI)-enabled network and pathway interrogation

To understand potential biological significance of proteins implicated by SPPB, 6MW, and the trial interventions, we performed two complementary pathway analytic approaches—a standard approach (Reactome) and an agentic AI-empowered approach. In each approach, we included proteins associated with SPPB or 6MW (at 10% FDR) in cross-sectional analyses <u>and</u> proteins that changed with intervention (at a nominal 5% level). We used standard approaches for Reactome-based pathway analysis^26,27^.

Our second approach integrated supercomputing, explainable AI, agentic AI with large language models (LLMs), and network analysis (additional details in **Supplemental Material**). We first constructed a 16-layer multiplex gene network to model multi-omic and tissue-specific mechanistic relationships using the *RWRtoolkit*^28^. This analysis required large-scale parallelization across hundreds of thousands of cores on the Summit and Andes supercomputers at the Oak Ridge National Laboratory (ORNL), enabling integration of over 16 distinct network evidence types spanning more than 64,539 (coding and non-coding) genes and 12,121,760 within/cross layer edges. Seven foundational layers were derived from HumanNet v3^29^, representing complementary evidence types: co-citation, co-expression, molecular pathways, gene interactions, gene neighborhood, phylogenetic relationships, and protein–protein interactions. We included transcriptional regulatory layers that were incorporated from public resources. The first was built from ENCODE^30,31^, representing experimentally derived transcription factor–target (TF–target) interactions across diverse human cell types. The second consisted of a TF–target network specific to human umbilical vein endothelial cells (HUVECs), derived from open-chromatin and eQTL mapping analyses relevant to vascular regulation^32^. Together, these layers capture both global and endothelial-specific transcriptional control networks important to vascular adaptation and remodeling. Additional predictive expression networks were constructed to include gene-gene relationships (see **Supplemental Material**). We used the resulting integrated networks in downstream MENTOR (Multiplex Embedding of Networks for Team-Based Omics Research) analyses to identify cross-layer mechanistic clades, functional convergence, and hierarchical biological themes^33,34^. MENTOR leverages the Random Walk with Restart (RWR) algorithm implemented in the *RWRtoolkit*^28^, which quantifies topological similarity among nodes within multiplex networks. Each protein was used as a seed node for RWR exploration, generating rank-ordered vectors of all nodes based on visitation probability. Each iteration of this high-resolution, distributed network traversal required approximately 3×10⁷ node-level probability computations, totaling roughly 10^9^ probability updates across proteins during network convergence. Using hierarchical clustering and an agentic AI-based interpretation framework (MENTOR-IA)^35^, an LLM-driven, retrieval-augmented generation (RAG) interpretation system that converts network embeddings into structured mechanistic narratives, we summarized the protein-network mapping to generate a mechanistic summary. In total, 11.8 million tokens were used as RAG input and 3.4 million tokens (≈480 pages of text) of output were generated by the LLMs. The MENTOR-IA framework was operated in a supervised, human-in-the-loop mode, where researchers reviewed and refined model outputs for accuracy, biological plausibility, and interpretability.

### Localizing targets in the physical function proteome at tissue- and single cell-level resolution through transcriptomics

To localize organs and cell types in which the physical function proteome was expressed, we interrogated tissue transcript expression (at a transcriptional level) of the same set of proteins subject to pathway analysis (above) in two major atlases: (1) the Genotype-Tissue Expression (GTEx) database^36^; (2) the Tabula sapiens human cell single-cell RNA-seq atlas^6^ (412,837 cells). Based on the strong result for skeletal muscle in these studies, the centrality of muscle in our other findings, and availability of serial biopsies, we also interrogated expression within skeletal muscle across additional human data: (1) Human Muscle Ageing Cell Atlas (HLMA)^8^ (210,445 snRNA-seq nuclei from 15 older and 7 younger donors); (2) a publicly available human aged skeletal muscle dataset (GEO identifier GSE167186; 29 older healthy, 19 younger healthy, and 24 sarcopenia subjects)^5^.

### Genomic approaches to tissue-specific liability to frailty and proteogenomic integration

#### Gene-level association

We next assessed whether this set of proteins (associated with physical function deficits) used in our pathway and tissue approaches had potentially causal effects on a model of deficit accumulation frailty^37^ via performing genetics-informed transcriptome-wide association studies (TWAS). TWAS integrates two types of data: (1) genome-wide association studies (GWAS; connects genetic variants to phenotype) and (2) expression quantitative trait loci data (eQTL; links genetic regulatory variants to gene expression in a specific tissue). TWAS tests whether genetically regulated expression (GReX) from eQTLs is associated with the phenotype. For this approach, we focused on potential tissues of origin prioritized by the proteome and single cell studies (skeletal muscle, heart left ventricle, and dorsolateral pre-frontal cortex [DPFC]). eQTL models in skeletal muscle and heart left ventricle were derived from GTEx (v8) data using well-established TWAS JTI methodology^38^. The models in DPFC were trained in PsychENCODE^38,39^ data. We leveraged the GWAS data from a multivariate study of the latent architecture across 30 frailty deficits (using the “general factor” from that work that encompassed all 30 deficits as a composite frailty index)^7^.

#### Integration of TWAS and intervention-induced proteomic changes

While the TWAS approaches nominate genes encoding the physical function proteome as involved in a model of deficit accumulation frailty, it does not address whether circulating proteins could serve as a liquid biomarker for this tissue-specific genetic susceptibility—a prelude to use of the physical function proteome as an outcome biomarker. We therefore developed a statistical framework, PRO-TWAS (PROteomics-intervention-informed Transcriptome Wide Association Study), that combines TWAS *summary statistics* (effect sizes of genes on frailty) and intervention proteomic (IP) statistics (protein log-fold changes). The goal of this approach was to evaluate whether genes identified through genetic prediction (TWAS) are consistent with potentially biological effects observed at the protein level under an intervention.

We formulated a TWAS-proteome gene prioritization score using only *summary* data (does not require access to individual-level data). We defined the gene-protein concordance statistic (GPCS):

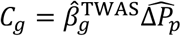

where 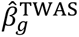 is the TWAS effect size estimate for the gene *g* and 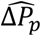 is the estimated intervention proteomic effect for the corresponding protein *p*. Given the null hypothesis of independence, the statistic *C_g_* has the following variance:

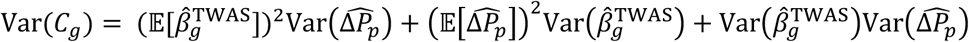

An unbiased estimate of Var(*C_g_*) is given by^40^:

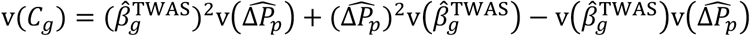

where 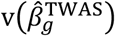 and 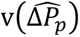 are unbiased estimates of 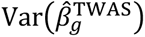 and 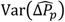 respectively.

We define the following Z-score:

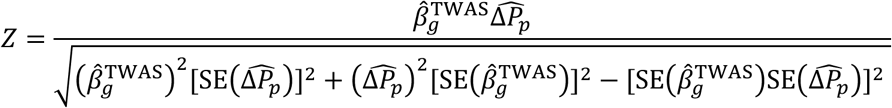

where 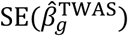 and 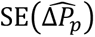 are the standard errors from TWAS and intervention-induced proteomic summary data, respectively, allowing hypothesis testing on the GPCS.

Next, we estimated the latent effect of the gene *g* on the protein *p* using the TWAS summary statistics and proteomic intervention statistics. Consider the following causal chain:

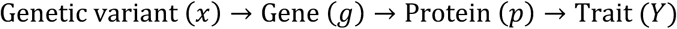

TWAS estimates the total effect *g* → *Y*, which we denoted as before by 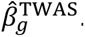. Mediator activation is given by *g* → *p*. However, we do not directly observe experimental manipulation of gene expression. Instead, we observe GWAS and eQTL-based TWAS effects (*x* → *Y* and *g* → *Y*, respectively) and proteomic changes under perturbation (perturbation → *p*). From these pieces of data, we need a method to extract the latent effect of the transcript *g* on the protein *p* (denoted by *γ*_*g* → *p*_). We posit that the intervention-induced protein change reflects the direction and magnitude of expression change for the corresponding gene. We can estimate *γ*_*g* → *p*_ as a function of GPCS *C_g_* and the TWAS summary statistics using errors-in-variables regression: is analogous to Mendelian Randomization 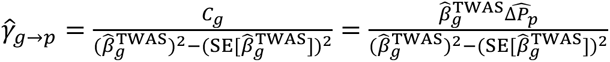 with TWAS as “instrument” and proteomic change as “exposure”, and the corrected estimator accounts for TWAS uncertainty.

We then developed a mediation framework to estimate the proportion of mediated effect (PME) using only summary data. We consider a latent mediation model in which the total genetic effect of gene expression on the trait is partially mediated by the protein level. Let *θ*_*g*_ be the latent total effect of gene *g* on the trait and *m*_*g*_ the proportion of the mediated effect (PME) by the protein. We posit the following model:

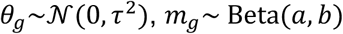

Thus, *m_g_* is Beta-distributed with shape parameters *a* > 0 and *b* > 0. PME is therefore given by:

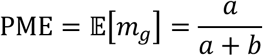

We estimate PME using a simulation-anchored method of moments. We use the observed correlation between the TWAS and proteomic intervention z-scores:

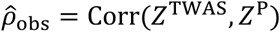

For each (*a*, *b*), we simulate latent z-scores under the model and calculate the expected correlation *ρ*_sim_(*a*, *b*). We estimate the optimal parameters 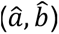 by minimizing the squared discrepancy:

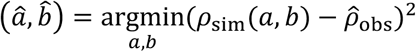

using the Nelder-Mead algorithm with initial values given by ((*Z*^P^)^2^, (*Z*^TWAS^)^2^). (*Z*^p^)^2^is the signal for the mediated path while (*Z*^TWAS^)^2^ is the signal for the direct path. The estimated PME is given by:

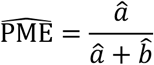

## RESULTS

### Characteristics of the study samples

The overall flow of our study is in **Figure 1**. This study included 126 REHAB-HF participants and 88 SECRET-II participants with HFpEF. REHAB-HF participants were older than SECRET-II participants (median age 73 vs. 68 years, respectively) with more physical impairment (median SPPB 6 vs. 10 points, median 6MW 157 vs. 376 meters in REHAB-HF and SECRET II, respectively; **Table 1**), approximately balanced by race. In contrast, participants in the UKB sample were younger (median age 58 years), predominantly White, with a low prevalence of frailty (58% with a Fried frailty score of 0; 39% with Fried frailty score of 1-2; and 3% with Fried frailty score ≥3; **Supplemental Table 2**).

**Table 1:** Characteristics of REHAB-HF and SECRET-II study populations. Continuous variables are reported as median (25^th^, 75^th^ percentiles). Categorical variables are reported as N (%). P values use Wilcoxon or Chi-square tests as appropriate and compare the control and intervention arms within each cohort. SPPB = Short Physical Performance Battery.

| Characteristic | REHAB-HF |  |  |  | SECRET II |  |  |  |
| --- | --- | --- | --- | --- | --- | --- | --- | --- |
|  | N | Control<br>N = 59 | Intervention<br>N = 67 | p-<br>value | N | Control<br>N = 44 | Intervention<br>N = 44 | p-<br>value |
| Age (years) | 126 | 73 (66, 78) | 73 (65, 78) | >0.9 | 88 | 67.3 (63.8, 71.8) | 69.0 (65.5, 72.4) | 0.14 |
| Women | 126 | 39 (66%) | 38 (57%) | 0.4 | 88 | 37 (84%) | 38 (86%) | >0.9 |
| Race | 126 |  |  | 0.5 | 88 |  |  | 0.8 |
| Black |  | 22 (37%) | 27 (40%) |  |  | 23 (52%) | 23 (52%) |  |
| White |  | 31 (53%) | 19 (43%) |  |  | 19 (43%) | 20 (45%) |  |
| Other |  | 6 (10%) | 2 (4.5%) |  |  | 2 (4.5%) | 1 (2.3%) |  |
| Body mass index (kg/m<sup>2</sup>) | 126 | 34 (26, 42) | 34 (29, 40) | 0.6 | 88 | 39.6 (36.2, 43.3) | 39.0 (35.8, 41.4) | 0.6 |
| Atrial fibrillation | 126 | 31 (53%) | 36 (54%) | >0.9 | 88 | 44 (100%) | 40 (91%) | 0.12 |
| Diabetes | 126 | 28 (47%) | 45 (67%) | 0.040 | 88 | 30 (68%) | 24 (55%) | 0.3 |
| Hypertension | 126 | 55 (93%) | 65 (97%) | 0.6 | 88 | 2 (4.5%) | 2 (4.5%) | >0.9 |
| Coronary artery disease | 126 | 18 (31%) | 20 (30%) | >0.9 | 87 | 40 (93%) | 43 (98%) | 0.6 |
| Current smoker | 126 | 6 (10%) | 7 (10%) | >0.9 | 88 | 17 (39%) | 22 (50%) | 0.4 |
| Total cholesterol (mg/dL) | 21 | 128 (115, 137) | 119 (114, 172) | 0.8 | 88 | 170 (152, 196) | 170 (138, 204) | 0.8 |
| Estimated glomerular filtration rate (mL/min/1.73m<sup>2</sup>) | 126 | 57 (32, 70) | 51 (38, 72) | 0.8 | 88 | 79 (64, 93) | 71 (58, 88) | 0.085 |
| SPPB, baseline | 126 | 5.00 (4.00, 7.50) | 6.00 (4.00, 8.00) | 0.8 | 88 | 10.00 (9.00, 11.00) | 9.00 (8.00, 11.00) | 0.078 |
| 6-minute walk distance (m), baseline | 121 | 151 (94, 233) | 168 (92, 257) | 0.7 | 88 | 388 (319, 410) | 375 (306, 406) | 0.4 |
| Gait speed (m/s), baseline | 126 | 0.54 (0.39, 0.69) | 0.54 (0.36, 0.76) | 0.8 | 88 | 0.84 (0.76, 0.98) | 0.92 (0.77, 1.07) | 0.5 |
| Right handgrip (kg), baseline | 124 | 22 (16, 26) | 24 (18, 30) | 0.3 | 85 | 24 (21, 32) | 26 (20, 30) | 0.6 |
| Left handgrip (kg), baseline | 121 | 20 (14, 26) | 22 (18, 28) | 0.3 | 86 | 25 (20, 30) | 24 (18, 28) | 0.4 |
| SPPB, follow-up | 109 | 7.0 (3.2, 8.0) | 8.0 (5.5, 10.0) | 0.017 | 79 | 11.00 (10.00, 11.00) | 11.00 (9.00, 12.00) | 0.5 |
| 6-minute walk distance (m), follow-up | 91 | 226 (121, 306) | 262 (178, 339) | 0.2 | 79 | 438 (396, 469) | 400 (356, 448) | 0.056 |

### Identifying proteins associated with physical function phenotypes

We identified 10 proteins associated with SPPB and 82 proteins associated with 6MW at 10% FDR in age- and sex-adjusted linear models (**Figure 2A**; **Supplemental Table 3**). Proteins associated with 6MW or SPPB prominently featured pathways with known physiologic domains relevant to muscle contraction (ACTA2, ACTN2, TTN^41^, MYL), substrate metabolism or mitochondrial integrity (ASAH2^42^, NDUFS6^43^, PLIN1^44^, CHCHD6^45^), myocyte repair and sarcopenia (CKAP4^46^, LIFR^47^, NOS1^48^, EDA2R^49^), cellular stress response and survival pathways (SUMO1^50,51^, ERBB2^52^, ANKRD2^53^), endothelial function and angiogenesis (EDN1, EGFL7^54^, ANGPT2), neuromuscular integrity and neural function (SORBS1^55^, NEFL1^56^), and immune activation (IL-6^57^, CD300E^58^, IL-15^59^, TNF pathway members), among others. Intracellular muscle contractile and mitochondrial proteins and essential for function were detectable in circulation and had an inverse association with physical function, consistent with previous studies suggesting sarcomere constituents or circulating mitochondrial-derived extracellular vesicles may reflect skeletal muscle injury^60^ and frailty^61,62^. Apart from systemic proteins in immunity, inflammation, and aging, we observed links between tissue inflammation and organ physiology relevant to physical function: for example, upregulation of EDA2R with aging has been shown to drive skeletal muscle loss and insulin resistance in part via NF-kB^63^, and NOS1—a key driver of both muscle regeneration^48^ and systemic inflammation^64^—was negatively associated with 6MW in our study. Proteins with a <u>positive</u> association with 6MW or SPPB (higher levels ∼ better physical function) had diverse roles, including synaptic regulation (C1QL2^65^), ceramide metabolism (ASAH2), cell survival and development (including in muscle, ERBB2^52^, LRRN1^66^), and intracellular lysosomal function (CTSF^67^), as well as a host of other proteins with roles in other organ systems (e.g., UMOD, kidney; EDDM3B, male reproductive) without previously described roles in frailty or function.

**Figure 2.**
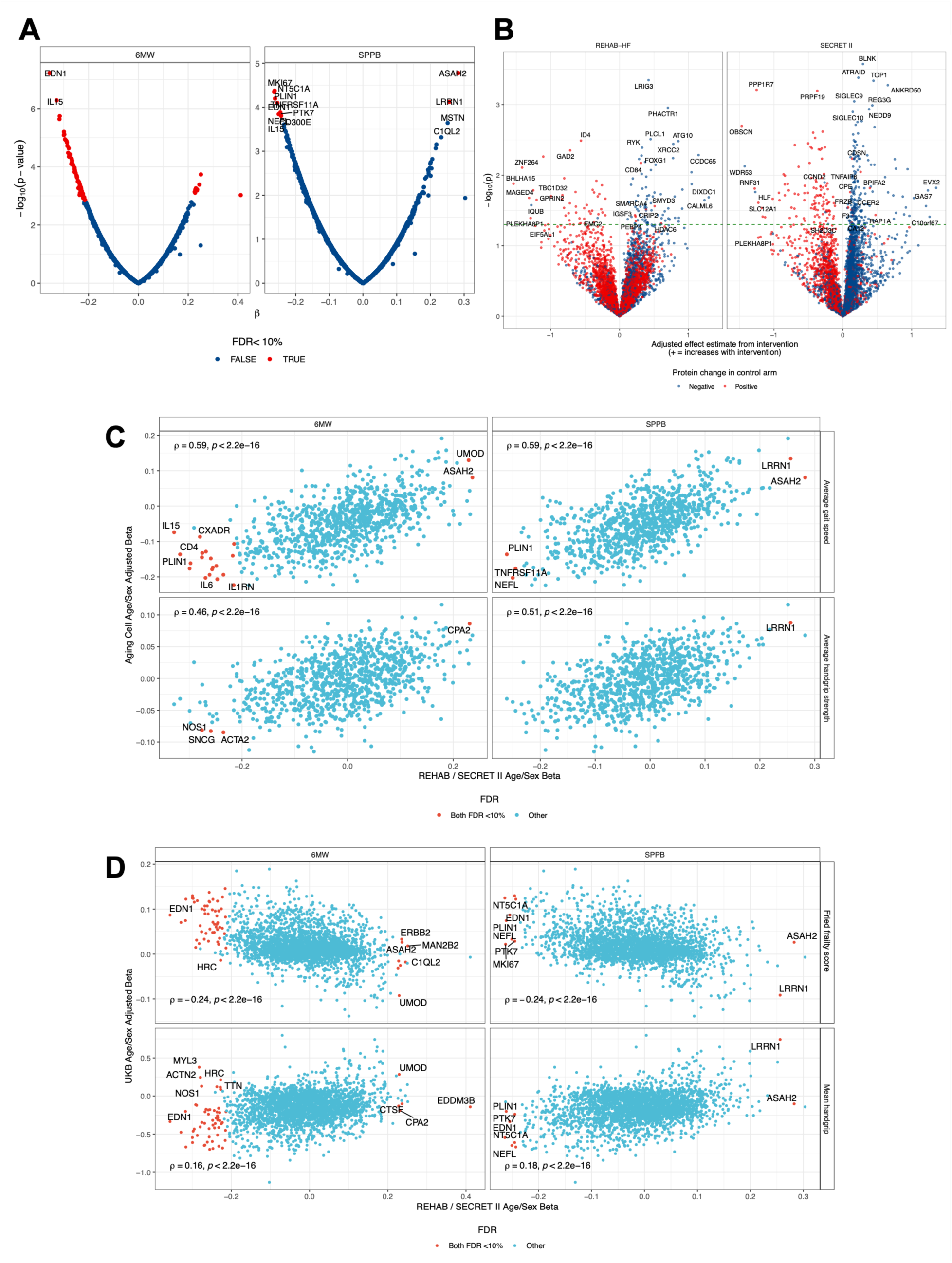
Proteomic architecture of physical function in heart failure with preserved ejection fraction. (A) Volcano plots showing age and sex adjusted linear models of SPPB and 6-minute walk. (B) Volcano plots showing estimated effect of intervention for linear models of post-intervention protein level, adjusted for baseline protein level. (C) Spearman correlations demonstrating concordance with linear models of SPPB and 6MW with frailty phenotypes measured in reference^4^. (D) Spearman correlations demonstrating concordance with linear models of SPPB and 6MW with frailty phenotypes measured in UK Biobank. 6MW = 6-minute walk distance. SPPB = Short Physical Performance Battery. FDR = false discovery rate, Benjamini-Hochberg method.

We observed a correlation ≈0.4-0.6 between protein-6MW or protein-SPPB regression effect sizes in REHAB-HF/SECRET-II with proteomic correlates of handgrip and gait speed previously reported by our group in older individuals with aortic stenosis (median age 83 years)^4^, a group exhibiting phenotypic similarity to the REHAB-HF/SECRET-II sample (**Figure 2C**). As expected, in a broader population with a lower prevalence of frailty (UKB), we noted a statistically significant but lower correlation (≈0.2) of our findings with proteomic correlates of Fried frailty score and handgrip in UKB (**Figure 2D**, **Supplemental Table 4**). Several of our strongest findings (by effect size) were directionality consistent across cohorts despite differences in functional measures utilized and differences in proteomic coverage, including proteins involved in inflammation (IL-15, IL-6, TNF family members), muscle contractile apparatus (ACTA2), substrate metabolism (ASAH2, PLIN1), myocyte repair and sarcopenia (CKAP4, NOS1), and neural structure (NEFL; all at 10% FDR).

### Identifying changes in the circulating proteome with rehabilitation

An advantage of integrating high-throughput molecular profiling within randomized controlled trials is the ability to unmask relevant molecules during perturbation that may be physiologically meaningful and dynamic^68–71^. While both REHAB-HF and SECRET-II included physical rehabilitation in their design, differences in delivery and ancillary treatment (e.g., caloric restriction) led us to stratify our analysis of proteomic changes after intervention by cohort (see **Methods**; **Figure 2B**; **Supplemental Table 5**). Among the top 20 targets in REHAB-HF (by significance level), we observed dynamic post-intervention alterations in targets related to muscle growth and maintenance, vascular aging, cell proliferation and senescence, autophagy, and metabolism (**Table 2**, **Supplemental Table 5**). While top targets from SECRET-II were distinct, they encompassed generally similar pathways. Of note, apart from proteins with established plausibility in aging, cardiometabolic, or muscle biology, we observed a host of other proteins with heretofore uncharacterized roles in aging, including proteins involved in cell renewal and cycling (PPP1R7, EPHB6, CCDC65), vesicle trafficking (ANKRD50), immune function (BLNK, CD84), and organ-specific biology (GARIN2, SPZ1).

**Table 2.** Curation of aging- or physical function-relevant proteins from top 20 targets different pre- versus post-intervention in SECRET-II and REHAB-HF.

| Protein | Cohort | Change with intervention | Potential mechanistic relevance |
| --- | --- | --- | --- |
| LRIG3 | REHAB | + | Increases BMP signaling<sup>110</sup>; involved in regulation of muscle growth and hypertrophy<sup>111</sup> |
| PHACTR1 |  | + | Decreases with age; involved in inflammation and atherogenesis<sup>112</sup> (vascular aging) |
| PLCL1 |  | + | Involved in bone and lean mass maintenance<sup>113</sup>; may regulate autophagy through activation of AMP kinase/mTOR pathways<sup>114</sup> |
| ATG10 |  | + | Involved in autophagy; deficiency can lead to cell senescence and death in non-human systems<sup>115</sup> |
| XRCC2 |  | + | DNA base repair |
| RYK2 |  | + | Cell proliferation; deficiency of RYK2 leads potentiate apoptosis<sup>116</sup> |
| GAD2 |  | - | Involved in GABAergic signaling; complex roles in aging; lower levels with age<sup>117</sup> may contribute to neuropsychiatric disorders. Involved in GABAergic signaling; complex roles in aging; lower levels with age<sup>117</sup> may contribute to neuropsychiatric disorders<sup>118</sup>; high levels impair insulin sensitivity<sup>119</sup> |
| FOXG1 |  | + | Activates autophagy and cell survival<sup>120</sup> |
| BTBD8 |  | + | Involved in intestinal inflammation; a loss of BTBD8 may reduce inflammation and increase cellular resilience in colitis<sup>121</sup> |
| CNGB3 |  | + | Involved in cone function in the retina, with expression related to improved visual function<sup>122</sup> |
| ZNF264 |  | - | Zinc finger protein; overexpression can induce aging-like phenotypes<sup>123</sup> |
| RICTOR |  | + | Central to mTORC2 signaling; reductions in expression can attenuate lifespan<sup>124</sup>, promote bone loss<sup>125</sup>, and alter muscle metabolism<sup>126</sup> |
| STARD13 |  | + | Deletion can lead to a defect in glucose-stimulated insulin secretion<sup>127</sup> |
| ADAMTS15 |  | + | Involved in matrix remodeling; reduced expression in fibroblasts from nonagenarians relative to children<sup>128</sup>Involved in matrix remodeling; reduced expression in fibroblasts from nonagenarians relative to children<sup>128</sup> |
| PRPF19 | SECRET-II | - | Central in DNA damage/repair response; can reduce aging and improve longevity in skin<sup>129</sup> |
| TOP1 |  | + | Topoisomerase 1; involved in torsional stress modulation during DNA processes; loss can contribute to neurodegeneration<sup>130</sup> |
| ATRAID |  | + | Context-dependent implications in bone health (with limited effects of osteoporosis treatments in its absence<sup>131</sup>) and senescence mechanisms<sup>132</sup> (tissue levels increasing with age)) and senescence mechanisms<sup>132</sup> (tissue levels increasing with age) |
| SIGLEC9/10 |  | + | Deletion of SIGLEC9 results in oxidative stress, DNA damage (accelerated aging)<sup>133</sup>; SIGLEC10 regulates immune responses in context of cancer (specifically targeting dendritic cells)<sup>134</sup> |
| REG3G |  | + | Anti-microbial protein; functions to enhance intestinal barrier and yield metabolic benefits through host-microbiome interaction<sup>135</sup>Anti-microbial protein; functions to enhance intestinal barrier and yield metabolic benefits through host-microbiome interaction<sup>135</sup> |
| NEDD9 |  | + | Loss of NEDD9 can lead to muscle atrophy in model systems<sup>136</sup>; polymorphisms in NEDD9 may be related to neurocognitive disorders<sup>137</sup> |
| BCL2L15 |  | + | Centrally involved in apoptosis, autophagy, and proliferation, with potential benefits on cell function with increased expression<sup>138</sup> |
| PGF |  | + | Deficiency linked to obesity and metabolic syndrome<sup>139</sup> |
| BAG3 |  | + | Linked to autophagy and age-related proteostasis<sup>140</sup> |
| OBSCN |  | - | Sarcomeric protein; deficiency in OBSCN can lead to aberrant cardiac calcium handling and alterations in cardiac cytoskeletal structure<sup>141</sup>Sarcomeric protein; deficiency in OBSCN can lead to aberrant cardiac calcium handling and alterations in cardiac cytoskeletal structure<sup>141</sup> |
| ENTPD2 |  | + | Time-dependent expression during neuroinflammation; inflammatory states decrease expression in the oligodendroglial cell line<sup>142</sup> |
| LTB |  | + | Lymphotoxin beta; linked to immune regulation (decrease in its receptor LTBR can contribute to senescence)<sup>143</sup> |

**Table 3.**
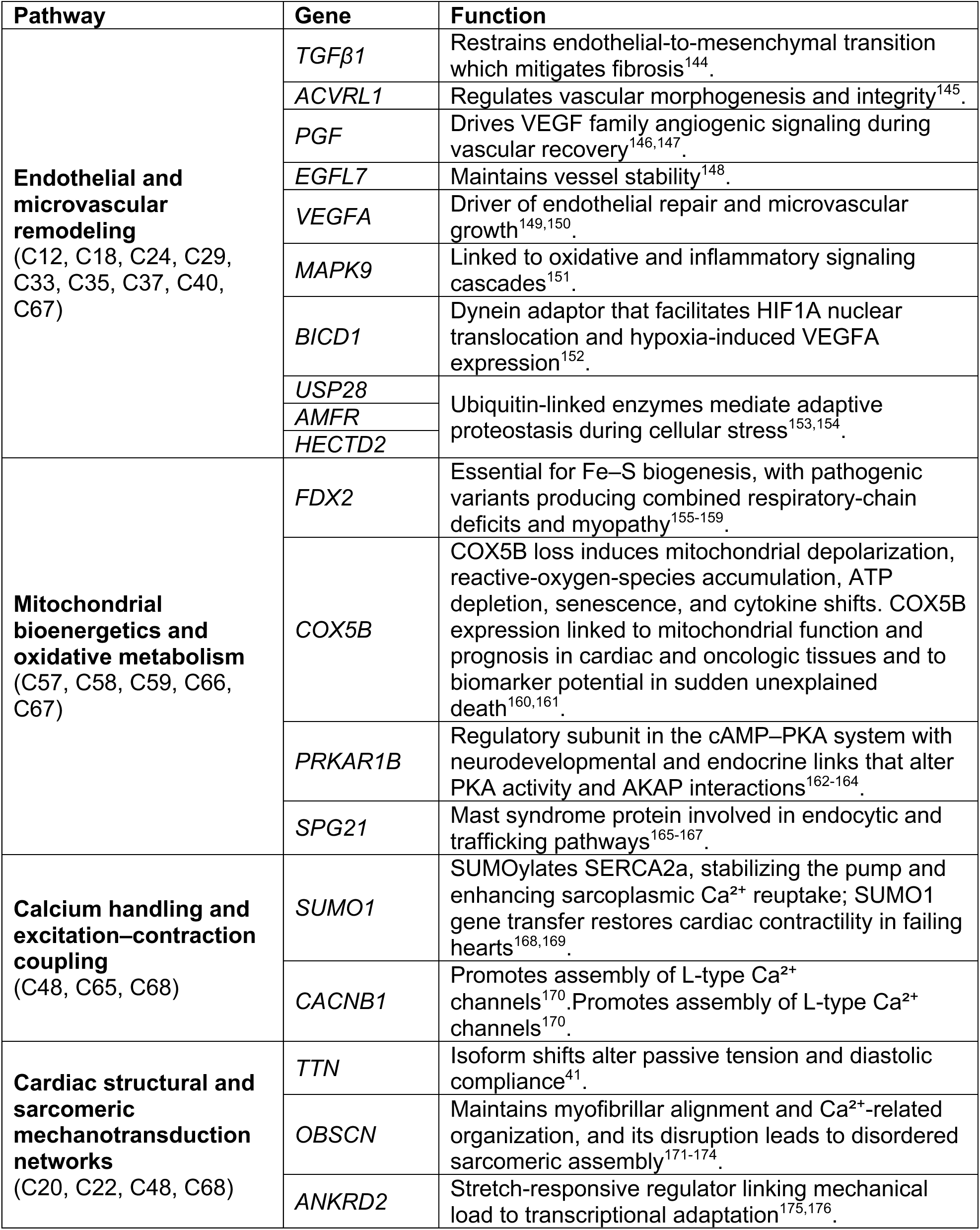
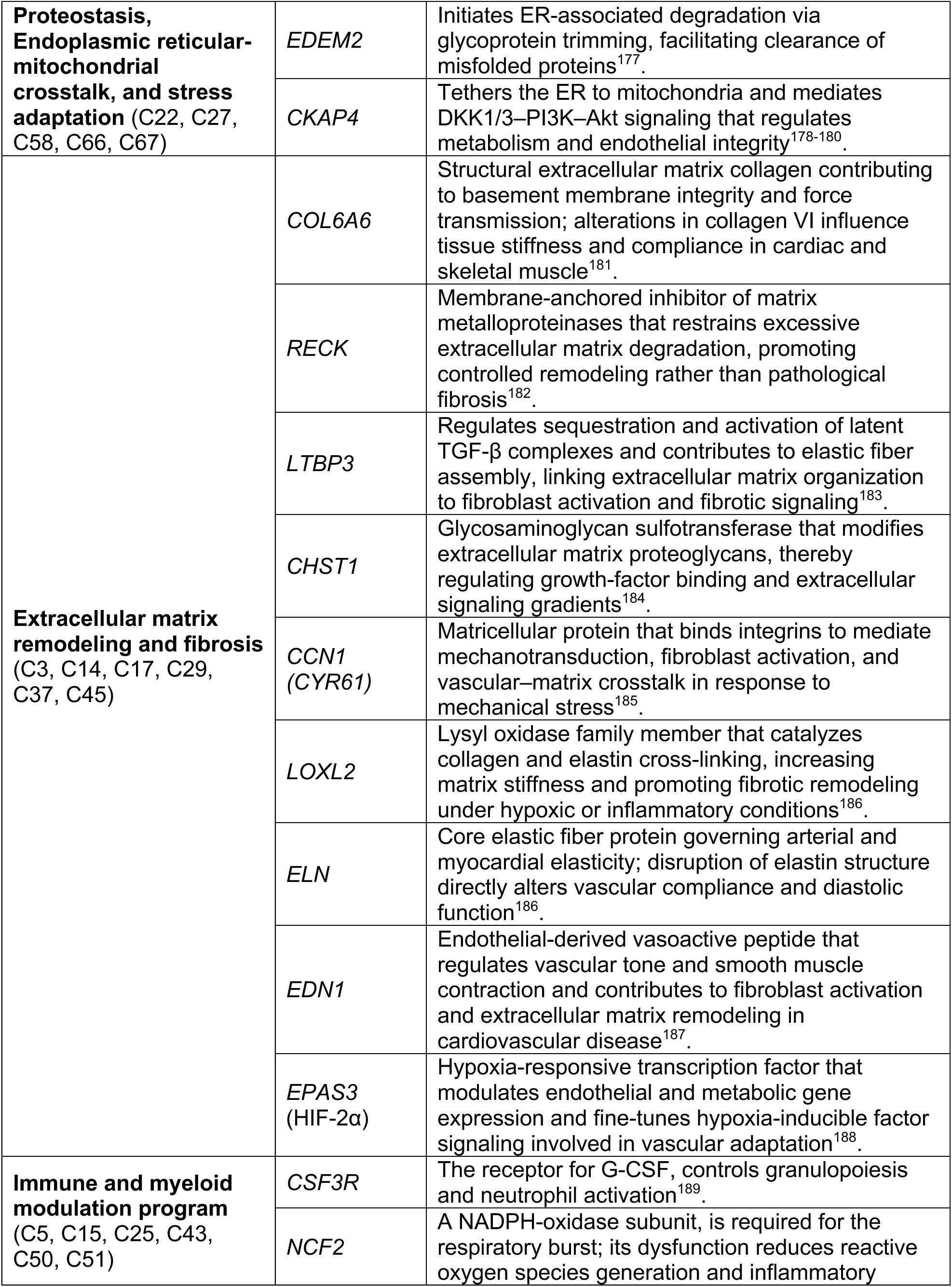
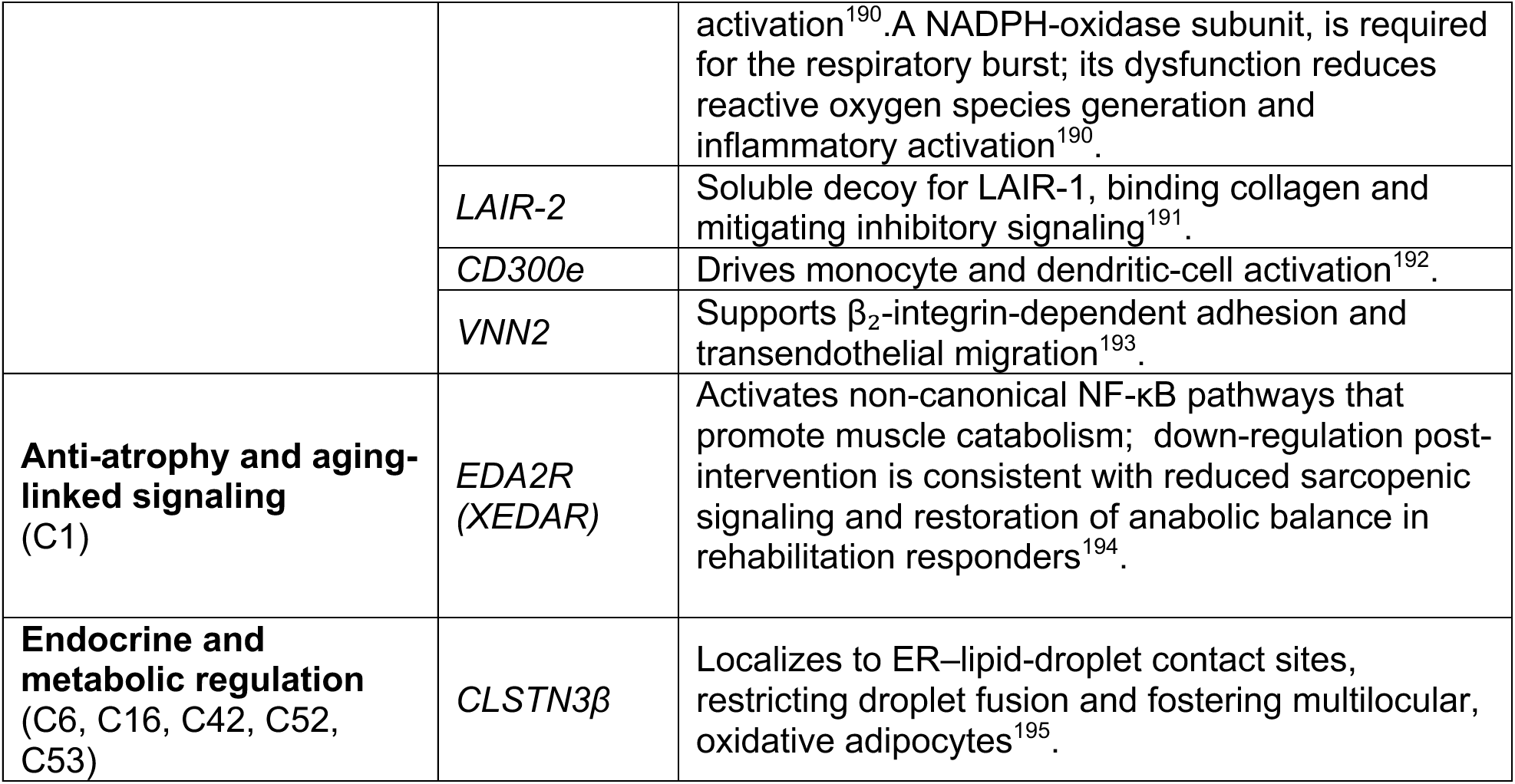
Multi-level, artificial intelligence-assisted curation of the physical function proteome.

### AI-powered network analysis identifies physiological pathways of physical function and frailty implicated by rehabilitation

To understand potential biological implications of these findings, we next mapped our findings to both a standard and an AI-driven networks analysis (see **Methods**). Both approaches utilized proteins prioritized from the following models (defined as a “physical *function proteome*”): (1) age-/sex-adjusted linear models of SPPB (at 10% FDR); (2) age-/sex-adjusted linear models of 6MW (at 10% FDR); (3) linear models estimating effect of physical rehabilitation on protein (stratified by trial cohort; at nominal P<0.05). **Figure 3** integrates molecular data from REHAB-HF and SECRET-II into a circular hierarchical framework from our AI-empowered analysis. The network topology prioritizes several key physiologic themes central to rehabilitation and HFpEF (specific genes for each theme in **Table 2**): (1) *endothelial and microvascular remodeling,* characterized by *TGFβ–ALK1–Notch–VEGF* signaling, angiogenic activation, nitric oxide bioavailability, and endothelial repair; (2) *mitochondrial bioenergetics and oxidative metabolism* (*FDX2*, *COX5B*); (3) *calcium handling and excitation–contraction coupling* (*SUMO1*, *CACNB1*, *TRPC4AP*). Other processes identified by our agentic AI-driven approach also recapitulated the multi-organ biology of HFpEF and physical function, including *cardiac structural and sarcomeric mechanotransduction networks* (contractile performance); *proteostasis, ER-mitochondrial crosstalk, and stress adaptation*; *fibrosis; immune and myeloid modulation*, among others. A more in-depth discussion of these themes is in the **Supplemental Material**. These themes expanded upon findings from standard pathway analysis (*Reactome*, **Supplemental Figure 3**). Together, these approaches strongly supported a multi-system physiologic program, implicating implicated by the physical function proteome across vascular function, mitochondrial energetics, and metabolism (systemic and muscle) as central to physical frailty in HFpEF.

**Figure 3.**
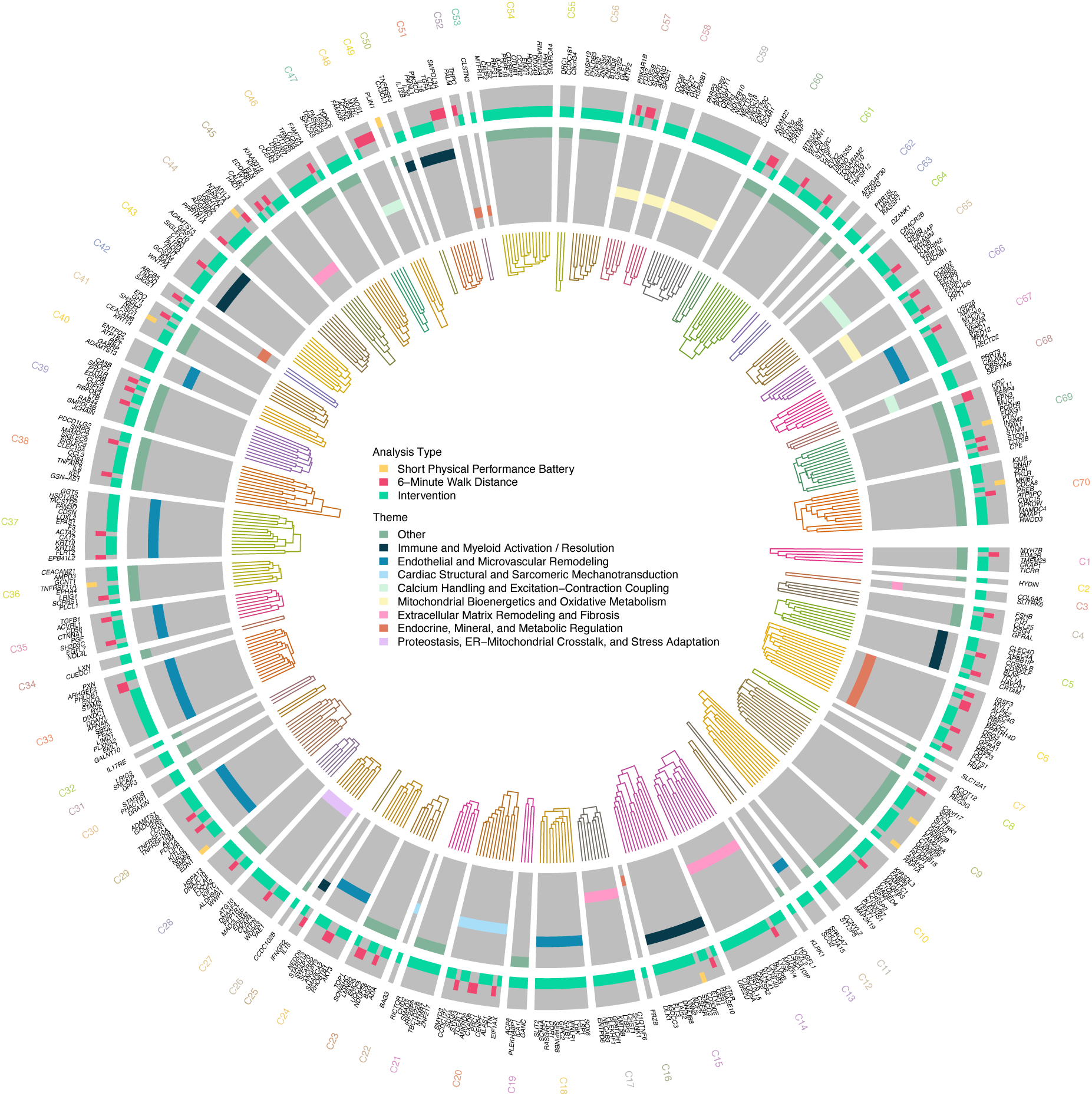
Artificial intelligence-assisted multiplex network topology of the HFpEF physical function proteome. Circular dendrogram showing hierarchical clustering of 475 proteins associated with baseline physical function (SPPB, 6MW) and rehabilitation-induced change. Proteins are organized based on similarity of their multiplex network connectivity profiles, derived from random-walk-with-restart embeddings across a 16-layer network integrating protein–protein interactions, transcriptional regulation, functional associations, and tissue-specific predictive expression networks. Branch length reflects topological distance, such that proteins that cluster closely share similar patterns of interaction across multiple biological layers, indicating greater mechanistic proximity. Colored tracks in the outer ring denote association with SPPB, 6MW, and/or pre- versus post-rehabilitation change. Inner annotation tracks indicate clade membership within the eight major mechanistic themes identified by MENTOR-IA. Together, the dendrogram provides a systems-level map of interconnected biological modules linking endothelial and microvascular remodeling; immune and myeloid activation/resolution; extracellular matrix remodeling and fibrosis; mitochondrial bioenergetics and oxidative metabolism; calcium handling and excitation–contraction coupling; cardiac structural and sarcomeric mechanotransduction; proteostasis, ER–mitochondrial crosstalk and stress adaptation; and endocrine and metabolic regulation.

### Tissue and cellular contexts of the physical function proteome

Across GTEx tissues, we observed a strong enrichment of genes encoding the physical function proteome (those associated with 6MW, SPPB, or changed during intervention) in three key tissues: skeletal muscle, heart, and cerebral cortex (**Figure 4A**). At a single cell level, analysis of 412,837 cells from the Tabula Sapiens human single-cell transcriptomic atlas^6^ were observed evidence of increased activity of these gene targets in heart and muscle, driven by 6MW/SPPB-associated targets (**Supplementary Figure 4**). Given the consistency of muscle tissue within both GTEx and Tabula sapiens and the availability of human aging muscle single cell data, we focused on aging muscle and sarcopenia (see **Methods**).

**Figure 4.**
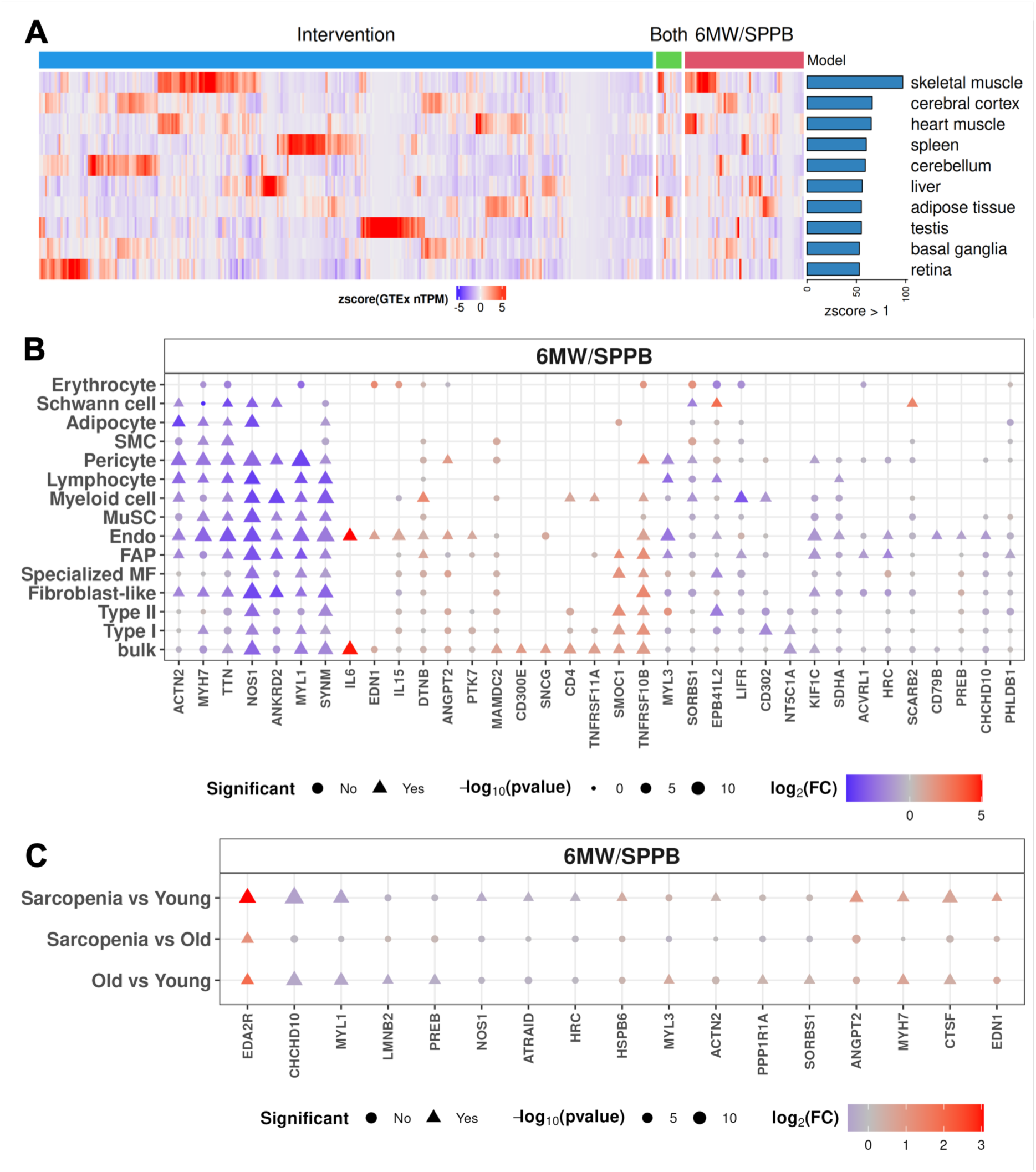
Tissue and cell-specific localization of the physical function proteome. (A) TPM values from GTEx were z-score normalized (centered and scaled) across tissues for each gene. The heatmap displays the top 10 tissues with the highest number of genes showing a z-score > 1. (B) Differential expression Human Muscle Ageing Cell atlas for targets associated with 6MW and SPPB. (C) Differential expression of targets associated with SPPB and 6MW across young, old, and sarcopenic lower limb samples (N=72). 6MW = 6-minute walk distance; SPPB = short physical performance battery.

We observed broad transcriptional differences by age in tissue transcripts implicated by the physical function proteome in the largest to-date published atlas of aging human muscle^8^, with expected cell-type or bulk tissue differential expression of key inflammatory (NOS1, TNF family members, IL6, CD300E) and muscle contractile genes (ACTN2, MYH5, TTN, MYL1) associated with SPPB or 6MW (**Figure 4B**). The proportion of significantly differentially expressed genes among those overlapping between 6MW/SPPB-associated proteins and expressed genes in each cell type reached up to 80%, with a median of approximately 30% (**Supplemental Figure 5A, 5B**). In addition, enrichment and dynamicity by age in skeletal muscle prompted us to focus on skeletal muscle to examine sarcopenia, a key driver of decreased physical function in HFpEF and with age.^5^ Using publicly available data on 72 lower limb muscle biopsies^5^, we found several genes encoding the 6MW/SPPB proteome that were differentially expressed in skeletal muscle by age (young versus older) and by sarcopenia status in older age: EDA2R (higher in sarcopenia/old compared to healthy/young), ANGPT2 (linked to muscle oxidative stress and inflammation that presage age-related muscle loss^72^; higher in sarcopenia compared to young healthy), and CHCHD10 (involved in mitochondrial integrity^73^, lower in sarcopenia and old healthy compared to young healthy), among others (**Figure 4C**). Muscle EDA2R and ANGPT2 exhibited directional consistency with our proteomic findings, while—analogous to our mitochondrial findings above—CHCHD10 was directionally distinct.

### Human genetics approaches link the physical function proteome to frailty

Next, to identify tissue-specific pathways pinpointed by the physical function proteome, we conducted a tissue-specific transcriptome-wide genetic association study (TWAS) to link the genes encoding physical function proteome to frailty (through a large published GWAS)^7^ in the three organs specified by our tissue- and single-cell studies (brain, heart, skeletal muscle). We selected a model of deficit accumulation frailty as our key outcome of interest for TWAS given its high prevalence and prognostic importance in HFpEF^1^ and its reversibility in REHAB-HF with rehabilitation^11^. We observed a substantial enrichment for gene-based associations with a measure of deficit-accumulation frailty (**Figure 5A**; labeled genes significant at 5% FDR). We observed the greatest number of significant targets in the brain, with implications in axonal development (*SLIT2*^74^), synaptic function (*GKAP1*), neuromuscular disorders (*SPG21*^75^), and neuroinflammation (*CD300LF*^76^). We identified one gene (*PCDH9*) in muscle, potentially linked to muscle performance^77^ and age-related peripheral nervous system function^78^. We observed correlation in significance in gene effects across tissues, with strongest relations in gene effects between heart and skeletal muscle and more modest effects with brain (**Supplementary Figure 6**), consistent with potentially shared mechanisms of genetic liability to frailty encoded by the physical function proteome across these key tissues.

**Figure 5.**
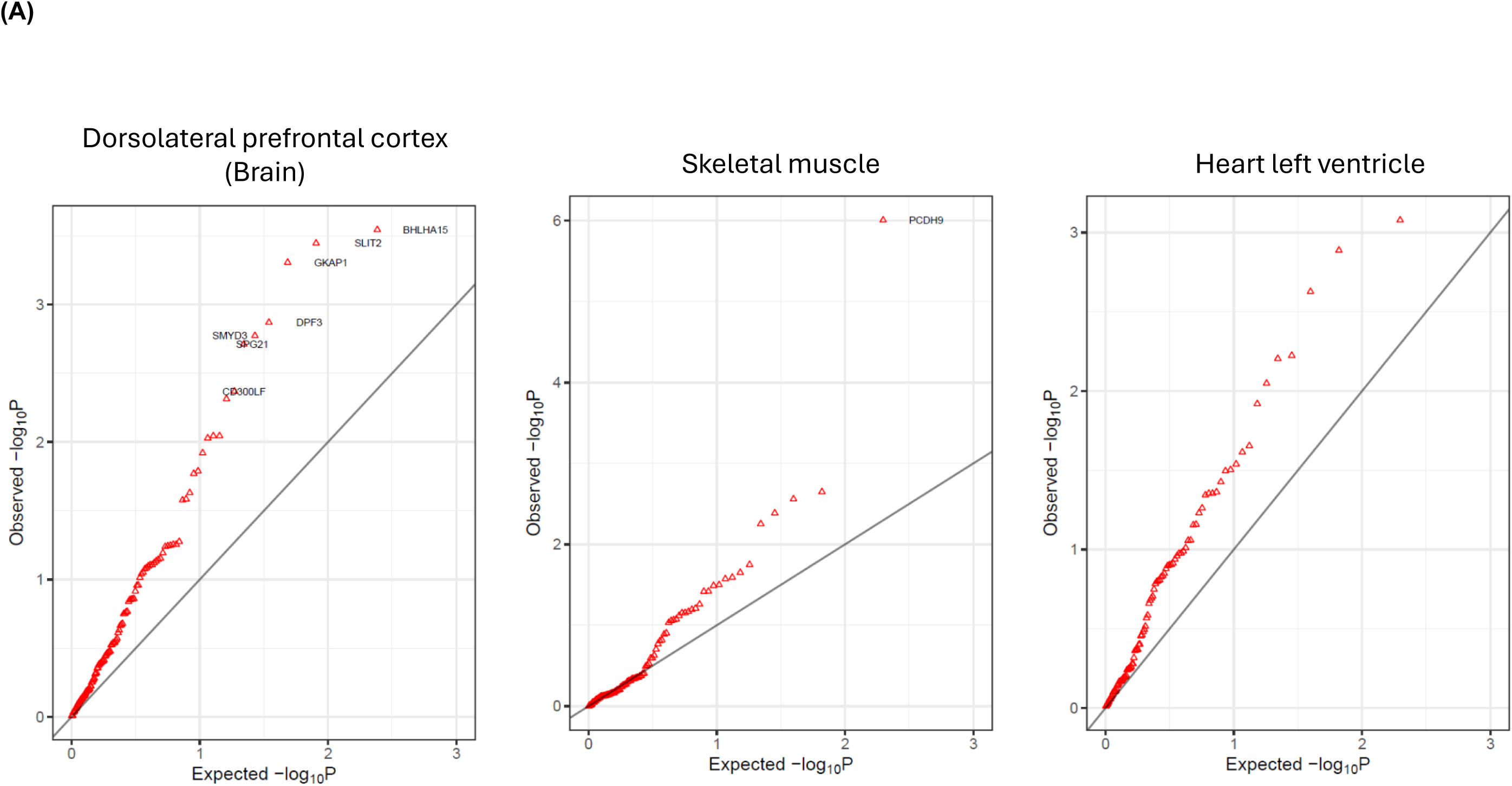
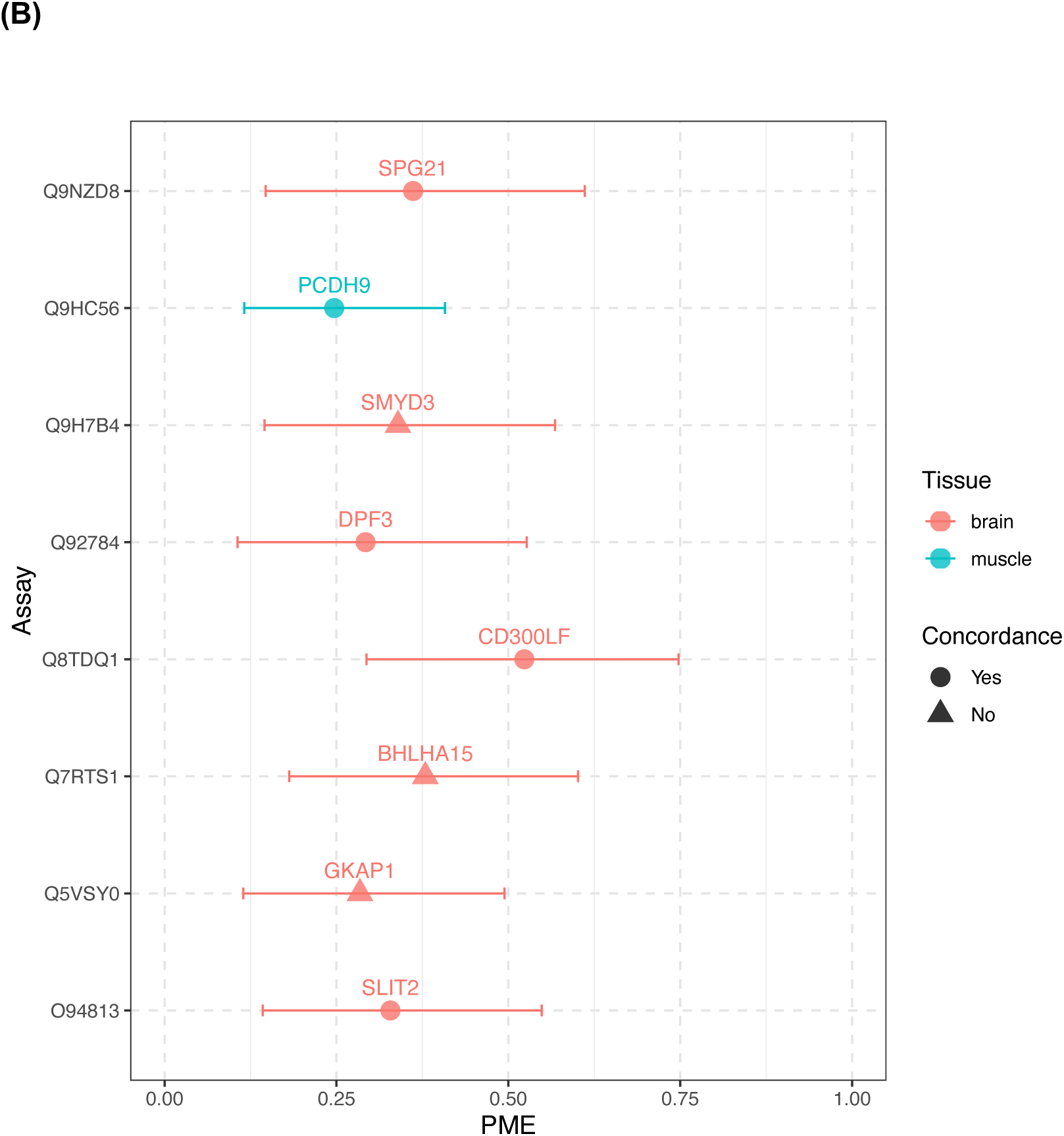
Human genomics maps the physical function proteome to tissue-specific liability for frailty. (A) Transcriptome-wide association study of frailty. For the proteins associated with SPPB, 6MW in REHAB-HF and SECRET-II at FDR<10% and proteins with changed expression after intervention in REHAB-HF or SECRET-II at a nominal P<0.05, we found a substantial enrichment for TWAS associations between genetically regulated gene expression (GReX) and a model of deficit-accumulation frailty. The TWAS analyses were conducted using gene expression models derived from dorsolateral prefrontal cortex (left), muscle (middle), and heart left ventricle (right). Departure from the diagonal line in the Q-Q plot indicates enrichment relative to null expectation. Labeled proteins were significant after multiple testing correction (at 5% FDR). (B) Mediation analysis (PRO-TWAS method) to quantify the extent by which the circulating protein mediates the effect of a tissue transcript (by human genetics) on frailty. Tissues included are brain and skeletal muscle, given they demonstrated significance in TWAS analysis (panel A). “Concordance” is “yes” if the direction of effect of the circulating protein on physical function (measured either as cross-sectional SPPB or 6MW or fold change with rehabilitation) is biologically consistent with TWAS effect. The estimates and bars reflect a point estimate with 95% CI.

Finally, to understand whether the circulating protein could serve as a liquid biomarker for tissue-specific genetic susceptibility, we quantified the extent to which the level of the circulating protein mediated the effect of its cognate tissue-specific transcript expression on frailty (PRO-TWAS method, see **Methods**). We found that the relationship between TWAS-implicated genes in brain and muscle with frailty were significantly mediated by the circulating protein level (**Figure 5B**; **Supplemental Table 6**). For example, the transcript *CD300LF* in brain implicated by our physical function proteome and TWAS approaches showed the highest mediated effect (0.52, 95% CI 0.29-0.75), consistent with its putative role as a systemic immune regulator relevant to cognitive decline, brain metabolism, senescence, and frailty^79^. Similarly, *PCDH9* (our top finding from muscle TWAS) exhibited a 25% mediated effect (95% CI 0.12-0.40). Collectively, these results suggest that select proteins within the physical function proteome may capture tissue-specific liability to physical function and frailty.

### Molecular signatures of physical rehabilitation are related to exercise capacity and associated with broad, clinical outcomes in aging and HFpEF in >26,000 individuals

Mediation results from human genetics and physiologically plausible pathway relevance of the physical function proteome suggested potential for proteomics to capture longitudinal risk of broad outcomes relevant to HFpEF and frailty. To this end, we identified a 28-protein and a 33-protein signature of SPPB and 6MW, respectively, across REHAB-HF and SECRET-II (**Figure 6A**, **Supplemental Tables 7, 8**). Multi-protein signatures demonstrated good model fit (Spearman ρ≈0.7 for both SPPB and 6MW; **Figure 6B-C**, **Supplemental Figure 7A-B**). <u>Lower</u> proteomic scores of physical function phenotypes were related to <u>greater</u> frailty (by Fried frailty score and handgrip) in UKB (recalibrated to proteins available in UKB, see **Methods**; **Figure 6G-H**, **Supplemental Figure 7E-F**) and <u>lower</u> peak VO_2_ as measured by cardiopulmonary exercise testing in SECRET-II (Spearman r≈0.3 for both SPPB and 6MW; **Supplemental Figure 7C-D**). Age and history of hypertension accounted for a significant proportion of variation in proteomic signatures (**Figure 6D**). Changes in each proteomic signature in REHAB-HF (but not SECRET-II) were related to changes in 6MW or SPPB pre- versus post-intervention (**Figure 6E-F**), potentially due to less frailty in SECRET-II, differences in mode of intervention type, and effect size of the intervention. We observed an association between pre-intervention proteomic signatures with future physical function status (SPPB standardized β=1.2, P=2.6×10^−5^; 6MW β=49.1, P=2.3×10^−5^), though non-significant after adjustment for pre-intervention (baseline) SPPB or 6MW. We did not observe significant interaction between either proteomic signature and rehabilitation on future physical function phenotypes.

**Figure 6.**
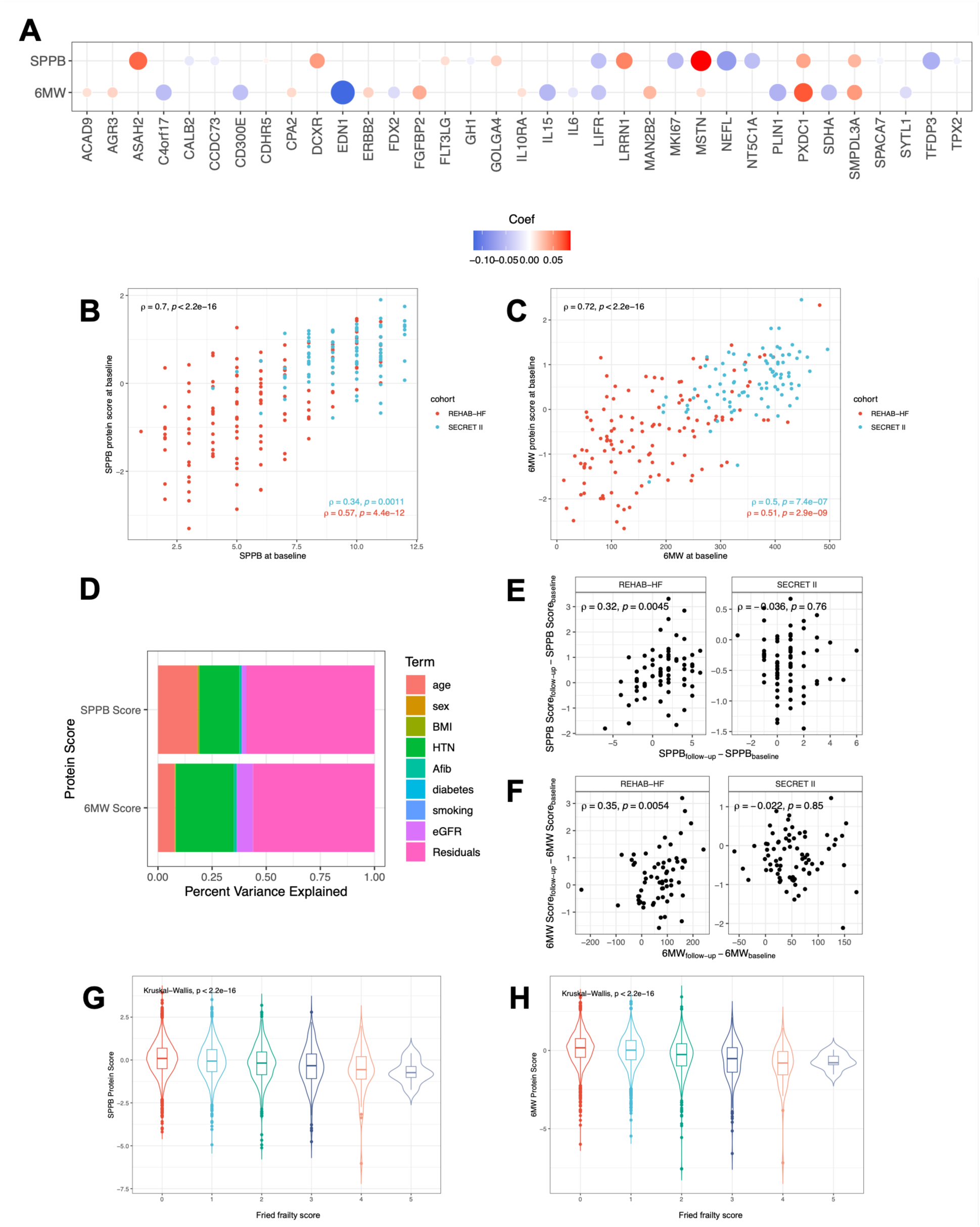
Proteomic signatures of physical function are dynamic and associated with frailty measures in UK Biobank. (A) Balloon plot of the LASSO model coefficients. For visualization, we selected the top 10 and bottom 10 (i.e., negative) coefficients for both SPPB and 6-minute walk with several proteins represented in both. (B, C) LASSO model fit in REHAB-HF and SECRET-II using the training data (baseline). (D) Type 1 ANOVA testing performed, which measures variance explained in order of variable appearance. Order of variables tested for variance was age, sex, body mass index (BMI), history of hypertension (HTN), history of atrial fibrillation (Afib), diabetes, smoking, and estimated glomerular filtration rate (eGFR). (E, F) Correlations of changes in protein scores with changes in the parent variables in REHAB-HF and SECRET. (G, H) Association of protein scores of SPPB and 6-minute walk with Fried Frailty Index in UK Biobank.

To assess the clinical relevance of the proteomics of physical function and rehabilitation, we assessed relation of proteomic scores and individual proteins related to 6MW or SPPB with multi-dimensional outcomes (reflecting the multi-organ nature of functional limitation in HFpEF) in ≈26,000 individuals in the UK Biobank. As noted in the **Methods**, given differences in proteomic platforms between REHAB-HF/SECRET-II and UKB, the multi-protein scores defined by LASSO were recalibrated for use in UKB. Individual proteins negatively associated with SPPB or 6MW in REHAB-HF/SECRET-II were generally associated with increased risk of clinical events (proteins related to increased physical function ∼ lower event hazard; **Figure 7**, **Supplemental Table 9**). Given differences in proteomic coverage between UK Biobank (Olink 3072) and REHAB-HF/SECRET-II (Olink HT), we recalibrated each proteomic signature (see **Methods**), with excellent performance in our derivation REHAB-HF/SECRET-II sample, suggesting transportability to UKB (ρ = 0.96 for 6MW, ρ = 0.97 for SPPB; **Supplemental Figure 8**, **Supplemental Table 10**). Over a median of 13.7 years of follow-up (25^th^-75^th^ percentile: 13.0-14.5 years, 2,762 mortality events), both SPPB and 6MW proteomic signatures were associated with all-cause mortality in fully adjusted models (SPPB: standardized hazard ratio HR 0.69 [0.66-0.72], P=1.13×10^−69^; 6MW HR 0.7 [0.67-0.73], P=9.27×10^−71^). Overall, both SPPB and 6MW signatures were associated with cardiovascular (ischemic heart disease, heart failure), neurologic (stroke, dementia), and fracture outcomes (**Figure 7**, **Supplemental Table 11**).

**Figure 7:**
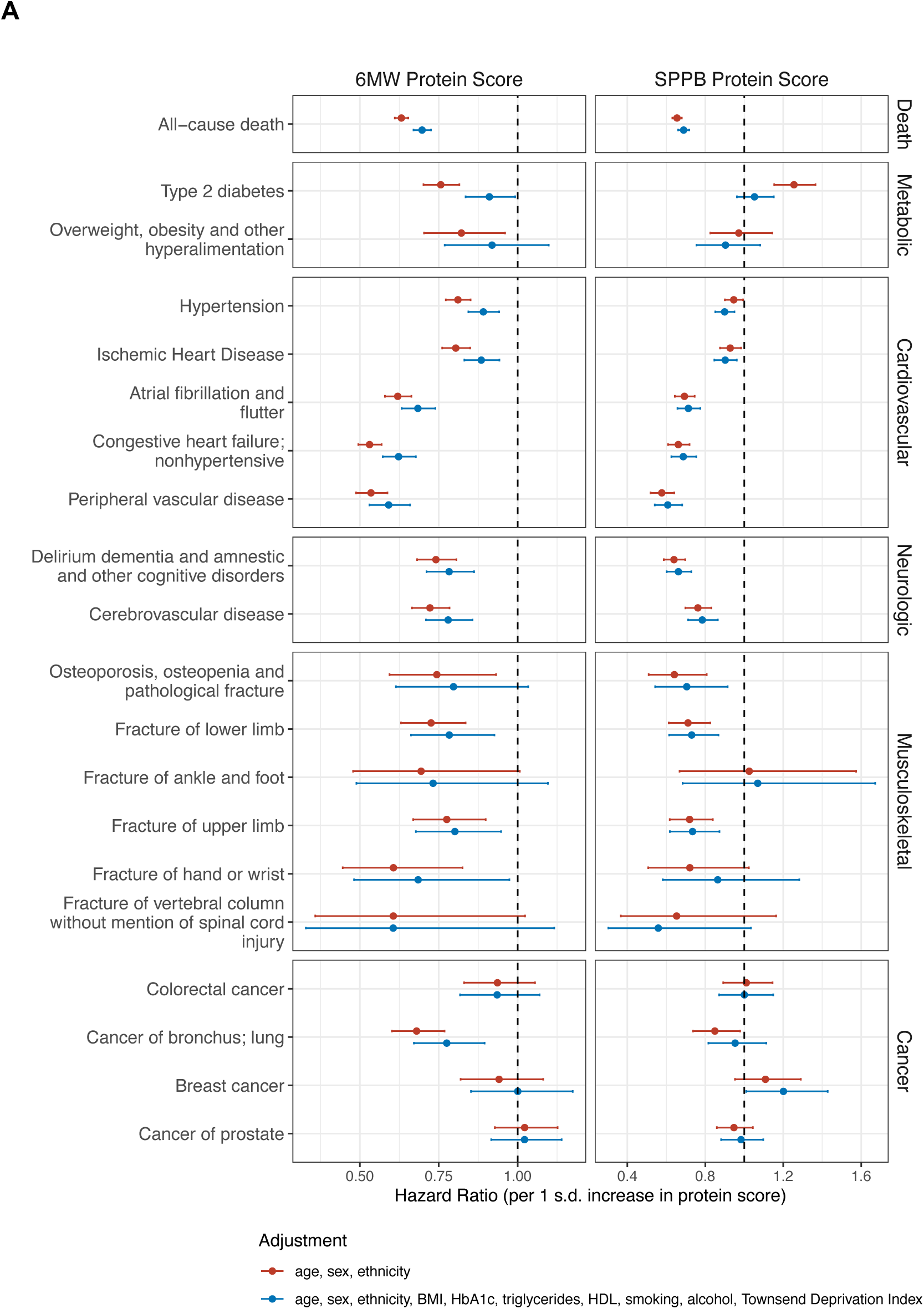
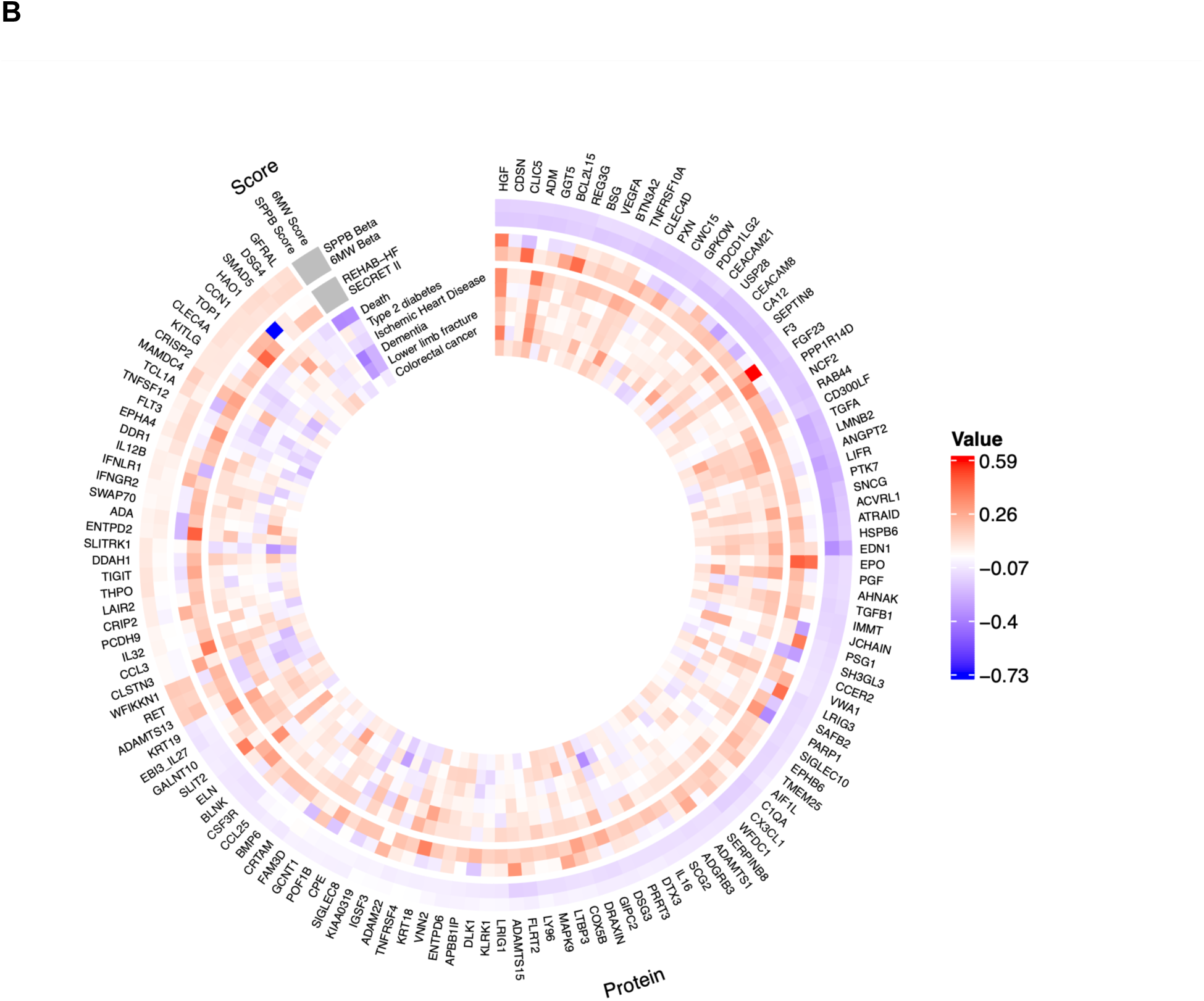
The physical function related proteome and its relation to clinical events. (A) Forest Plot showing adjusted, standardized hazard ratios of multi-protein signatures with clinical outcomes in UK Biobank. (B) Summary heatmap showing the concordance between proteomic relations with SPPB and 6MW (SPPB Beta, 6MW Beta), how protein levels change with intervention in REHAB-HF and SECRET-II (REHAB-HF, SECRET II), and their relations with clinical outcomes in UK Biobank.

## DISCUSSION

Functional limitation is a cardinal symptom of HFpEF. Building on evidence from HF-ACTION (a HFrEF cohort)^80^, several studies have attempted to improve physical function and quality of life through rehabilitation; they have consistently demonstrated improved functional measures and may potentially influence morbidity and mortality^3,11,13,81–83^. Despite numerous large, adequately powered, multi-center randomized controlled trials testing dozens of promising agents in HFpEF, nearly all trials have had negative results on their primary outcomes, such that few proven treatments are available for this large and growing, high risk population. Accordingly, there is an urgent need to identify new molecular targets for intervention in HFpEF. Here, we quantified a broad circulating proteome in two randomized clinical trials of rehabilitation and exercise training in HFpEF (REHAB-HF, SECRET-II) to pinpoint mechanisms responsible for the clinical effect of rehabilitation. Using both curated and large-scale AI-based methods, we found that proteins dynamic with intervention or associated with physical function in the HFpEF trials (the “physical function proteome”) implicated pathways of skeletal muscle function, cellular stress response and senescence, endothelial and neuromuscular function, immune activation, and mitochondrial integrity and metabolism. Furthermore, we observed high expression of genes encoding the physical function proteome in three major tissues—brain, skeletal muscle, and cardiac—with several key genes dynamic with age and sarcopenia in human muscle at single cell resolution (*EDA2R*, *ANGPT2*, *CHCHD10*). Tissue-specific human genetic approaches specified several genes from the physical function proteome with potential causal implications on frailty in brain (*SLIT2, GKAP1, SPG21, CD300LF*) and muscle (*PCDH9*). Moreover, several proteins appeared to mediate the genetic effect of tissue-specific gene expression on frailty, implicating the physical function proteome as a potentially functional biomarker of disease. As a consequence, we defined multi-protein signatures of physical function, demonstrating their strong association with mortality and cardiovascular, neurologic, and musculoskeletal (e.g., fracture) outcomes across >26,000 adults. By linking HFpEF randomized trials, multi-level AI-driven pathway interrogation, genomics, and longitudinal studies, these results identify potentially causal multi-organ mechanisms underlying physical function in HFpEF with implications on long-term outcome, precision targeting of rehabilitation intervention, and potential therapeutic development.

The use of circulating proteomics to implicate HFpEF pathobiology is emerging^84–90^, with most studies attempting to sub-phenotype HFpEF^84–86,88,89,91^ and fewer utilizing longitudinal study design to identify dynamicity with therapy (e.g., spironolactone^87^, SGLT2 inhibition^92^). A key recurrent theme across both types of studies in HFpEF is the importance of a fibrotic-inflammatory proteome on cardiac performance^85^, functional capacity^84^, and therapeutic response^87,91^. In one of the largest to-date proteomic studies of functional capacity in HFpEF (VITALITY-HFpEF), deFilippi and colleagues performed unsupervised clustering across 763 individuals with HFpEF with 368 proteins, identifying an inflammatory proteome linked to frailty, patient-centered outcome, and 6MW (e.g., TNFRSF11A, also found in our study)^84^. By comparison with our cohort, relation of proteomic modules to frailty and 6MW distance were more modest^84^ (correlation in range ≈0.1-0.2), and REHAB-HF was more functionally impaired (by 6MW; VITALITY, median 300 m). Our study extends prior work in HFpEF by studying the proteomics of a readily deployable, proven intervention and prognostic phenotypes (rehabilitation and physical function), with integration across tissue physiology and genomics, broad populations across cardiovascular disease liability (UKB, AS), and outcome. We observed concordance of protein-phenotype association across the spectrum of risk (UKB and AS), implicating putative pathways within HFpEF and aging: structural integrity, turnover, and maintenance of the sarcomere, mitochondrial function, cellular stress responses, endothelial function, and immune and neural homeostasis^93–95^. Importantly, many associations were *negative*, reflecting an increased protein abundance in circulation related to a lower functional phenotype, consistent with proposed mechanisms of cellular leak (including extracellular vesicle release^61,62^) in injured, degenerating, or atrophied myocytes^96–100^. Higher levels of *circulating* NDUFS6 or CHCHD10 (both *mitochondrial* proteins essential for energy generation^43,101^) and PLIN1 (involved in lipid metabolism^44^)—both intracellular proteins not usually in plasma—were also associated with decreased functional capacity. The prioritization of non-cardiac targets in our study highlights the importance of <u>peripheral</u> mechanisms in HFpEF, consistent with clinical trial evidence supporting the use of metabolic therapy (like SGLT2i) in HFpEF^102,103^.

In addition to examining the pre-intervention proteome, a key innovation of our approach was studying changes with a proven randomized intervention in a high-risk HFpEF population. While we were limited by sample sizes inherent to careful conduct of rehabilitation studies in older individuals with HFpEF, our approach leveraged a multi-stage, integrated approach to discovery, with broad protein-phenotype and protein-rehabilitation change analyses (across >5000 proteins) in our randomized trials, with three integrated studies to support biological implication and generalizability: (1) a large-scale, agentic AI-assisted supercomputing approach; (2) tissue and genomic approaches at single cell resolution in key tissues; (3) large-scale prognostic studies.

First, our novel AI-driven approach integrated 16 distinct evidence types (co-citation, co-expression, molecular pathways, gene interactions, gene neighborhood, phylogenetic relationships, protein-protein interaction, transcriptional regulatory networks) in an interactive process requiring 10^9^ real-time probability updates, followed by agentic AI large language model-based summarization. While both AI-based and standard pathway approaches (e.g., Reactome^104^) can incorporate curated resources, the agentic AI-based interpretation enables real-time dynamic learning, adaptation, and construction of interpretable, annotated networks with biological plausibility. Indeed, based on our physical function proteome, we identified both cell type-specific (endothelial and microvascular remodeling, myocyte-based processes) and more general cellular physiologies (e.g., mitochondrial metabolism, immune modulation, proteostasis) not necessarily easily discernable by other approaches^93–95,105^ but consistent with published model studies in HFpEF^106^.

These findings complemented tissue-based and single cell studies (our second approach), which suggested the brain, heart, and skeletal muscle as prime organs of high transcript expression of the physical function proteome—consistent with the AI-based pathway approach pointing to mitochondria, vascular, and immune function. Indeed, several transcripts identified in relation with SPPB or 6MW and prioritized by the agentic AI approach exhibited differences in expression with age and with sarcopenia in *skeletal muscle* (e.g., EDA2R, linked to age-related sarcopenia^49^). Localization to brain, skeletal and heart muscle at a transcriptional level allowed us to use standard and novel human genetic approaches to link the physical function proteome to multi-factorial frailty, demonstrating significant enrichment for transcript-level associations with frailty for targets from our physical function proteome, particularly in the brain and muscle. Using a new analytic approach (PRO-TWAS), we concluded that a significant fraction of tissue-specific genetic liability to frailty may be mediated by select circulating proteins relevant to muscle metabolism and synaptic function. While our reference genome-wide association studies used here were not in HFpEF, these results amplify the multi-organ relevance of physical function broadly, further suggesting a role for the circulating proteome *defined in HFpEF* as a potential functional biomarker of frailty.

This aggregate associative, biological, and genetic data finally drove us to construct and test a circulating proteomic signature of our key functional phenotypes (SPPB and 6MW) against multi-system outcomes relevant to HFpEF as an index of clinical relevance. With clinically translatable multi-protein scores, we observed a nearly 30-40% reduction in multi-system disease risk across >26,000 individuals in the UK Biobank: protective associations with all-cause and cardiovascular mortality, diabetes, dementia and cerebrovascular disorders, and fracture or osteopenia. The observed effects on outcome were higher than recently reported polygenic predictors of frailty^107^ and have comparable effects on outcome with fewer protein constituents than other signatures of physical performance^108^. The dynamicity of these multi-protein scores within REHAB-HF underscores their ability to monitor therapeutic response and may be informative in personalizing rehabilitation prescriptions. Importantly, identification of multi-system impact of a functional-derived signature on bone- and brain-related outcomes is consistent with our human genetic results and extant literature linking gait and neurocognitive performance^109^. These findings demonstrate potential of an individual’s physical function proteome to implicate specific extra-cardiac multi-morbidity.

Our study represents the largest, to-date evaluation of physical function in HFpEF with a rehabilitation intervention, deploying a multi-level framework across randomized trials, agentic AI-powered biological curation, tissue and genetic approaches and large human population validation. We recognize that the sample size in the two randomized clinical trials here is limited relative to our measured proteome, a challenge for any multi-omics study in phenotypically intensive randomized studies in patient subsets (relative to epidemiologic settings). Nevertheless, a key innovation of our study is the use of multiple layers of evidence, which added to generalizability of our findings in REHAB-HF and SECRET-II. Specifically, the physical function proteome (1) displayed concordant effects with frailty and functional endpoints in multiple settings (AS, UK Biobank, post-intervention phenotypes in the trials); (2) exhibited biological plausibility through curated and agentic AI-based network interrogation of physiology; (3) was strongly linked to phenotypes and outcomes in large cohorts (UK Biobank); (4) exhibited human genetic evidence linking protein expression to frailty. In addition, the studies here were performed in NIH-funded clinical trials of rehabilitation in HFpEF with proven prognostic phenotypes and clinical effect. While larger, more definitive sample sizes with homogeneity in rehabilitation intervention are currently underway (REHAB-HFpEF, clinical trials identifier NCT05525663), multi-layer clinical, biological, and genetic support lends validity to the result.

In conclusion, in this study of rehabilitation in HFpEF, we incorporated (1) high-throughput proteomic profiling within interventional clinical trials, (2) novel AI-assisted network analysis methods, and (3) innovative human genetics approaches to identify molecular mediators of frailty and physical function in older adults with HFpEF. We identified biological networks modified by rehabilitation including endothelial remodeling, mitochondrial metabolism, calcium handling, and immune modulation. Prioritized targets localized to heart, skeletal muscle and brain tissues were implicated by tissue-specific transcriptome-wide association studies in the pathogenesis of frailty. In addition, select circulating proteins were implicated as mediating the effect of tissue transcription on frailty. Multi-protein signatures of physical function correlated with functional changes observed with rehabilitation and were associated with heart failure and multi-dimensional clinical outcomes. These findings delineate a multi-system molecular program underlying physical function and rehabilitation response in HFpEF, offering insights into potential therapeutic targets to enhance physical function in HFpEF.

## Data Availability

All data produced in the present study are available upon reasonable request to the authors.

## ACKNOWLEDGEMENTS

This study was funded by the Department of War (HT94252410035 to ASP) and National Institutes of Health (R21AG086679 to MN and RVS). Dr. Perry is supported by the NIH (K23HL179316). This multiplex network and AI methods used in this manuscript were supported by NIH grants DA051908 (MP, ML, KS, AT, AV, BN, DAJ), DA051913 (MP, ML, KS, AT, AV, BN, DAJ), and DA054071 (MP, ML, KS, AT, AV, BN, DAJ). MN is supported by NIH grants R01HL156975, R01HL131029, and R01HL175323. This research used resources of the Oak Ridge Leadership Computing Facility at the Oak Ridge National Laboratory, which is supported by the Office of Science of the U.S. Department of Energy under Contract No. DE-AC05-00OR22725. This manuscript has been co-authored by UT-Battelle, LLC under Contract No. DE-AC05-00OR22725 with the U.S. Department of Energy. The United States Government retains and the publisher, by accepting the article for publication, acknowledges that the United States Government retains a non-exclusive, paid-up, irrevocable, world-wide license to publish or reproduce the published form of this manuscript, or allow others to do so, for United States Government purposes. The Department of Energy will provide public access to these results of federally sponsored research in accordance with the DOE Public Access Plan (http://energy.gov/downloads/doe-public-access-plan). The REHAB-HF and SECRET-II trials were supported by National Institutes of Health grants R01AG045551 and R01AG18915; they were also supported in part by grants R01AG078153, P30AG021332, U24AG059624, U01 AG076928, U01HL160272 and P30AG028716 (AMP). The trials were also supported in part by the Kermit Glenn Phillips II Chair in Cardiovascular Medicine and by the Oristano Family Fund at Wake Forest University School of Medicine.

