## Supplemental Materials for "A multi-layered approach to elucidate mechanisms of physical function in response to rehabilitation in heart failure with preserved ejection fraction"

### SUPPLEMENTAL METHODS

#### Artificial intelligence-enabled multiplex network analysis

**Molecular network assembly:** We constructed a 16-layer multiplex gene network to model cross-omic and tissue-specific mechanistic relationships relevant to rehabilitation biology in HFpEF using the *RWRtoolkit*[1]. This analysis required large-scale parallelization across hundreds of thousands of cores on the Summit and Andes supercomputers at the Oak Ridge National Laboratory (ORNL), enabling integration of over 16 distinct network evidence types spanning more than 64,539 (coding and non-coding) genes and 12,121,760 within/cross layer edges. Network assembly followed the *RWR_make_multiplex* workflow, which integrates heterogeneous biological evidence into a unified, multi-layer graph structure suitable for random-walk-based inference. Seven foundational layers were derived from HumanNet v3[2], representing complementary evidence types: co-citation, co-expression, molecular pathways, gene interactions, gene neighborhood, phylogenetic relationships, and protein–protein interactions. We included transcriptional regulatory layers that were incorporated from public resources. The first was built from ENCODE[3,4], representing experimentally derived transcription factor–target (TF–target) interactions across diverse human cell types. The second consisted of a TF–target network specific to human umbilical vein endothelial cells (HUVECs), derived from open-chromatin and eQTL mapping analyses relevant to vascular regulation[5]. Together, these layers capture both global and endothelial-specific transcriptional control networks important to vascular adaptation and remodeling.

Finally, to complement these literature- and experiment-based sources, we generated Predictive Expression Networks (PENs) using our iterative Random Forest–LOOP (iRF-LOOP) algorithm[6] applied to bulk-tissue gene expression data from the Genotype-Tissue Expression (GTEx) project[7]. PEN construction required extensive compute resources, as iRF-LOOP performs iterative feature selection and conditional dependency modeling at genome scale. Using the Summit supercomputer at ORNL, we generated seven high-resolution, tissue-specific

PENs capturing predictive gene–gene relationships across cardiometabolic and systemic organs (aorta, atrial appendage, left ventricle, coronary artery, whole blood plus EBV-transformed lymphocytes, lung, and liver). Each PEN encodes tissue-specific predictive dependencies among genes, thus capturing context-relevant transcriptional relationships that complement the global HumanNet and TF–target layers. All layers were normalized and integrated using the multiplex assembly module of *RWRtoolkit* on the Andes supercomputer at ORNL, resulting in a cohesive multi-evidence, multi-tissue framework. The resulting integrated networks served as the foundation for downstream MENTOR (Multiplex Embedding of Networks for Team-Based Omics Research) analyses to identify cross-layer mechanistic clades, functional convergence, and hierarchical biological themes.

We applied this MENTOR framework[8,9] to proteins prioritized from the above analyses relating circulating proteins to pre-intervention SPPB and 6MW (see “Identifying proteins associated with pre-intervention SPPB and 6MW” in **Methods**) and the impact of rehabilitation intervention on protein levels (see “Examining changes in the proteome with intervention” in **Methods**). The union of all significantly differentially expressed proteins was integrated into MENTOR to characterize functional and adaptive molecular phenotypes, with corresponding β coefficients visualized as heatmap tracks and cohort origin (REHAB-HF, SECRET-II, or both) indicated for each protein. Clades participating in 8 different mechanistic themes identified by MENTOR-IA (see below) were also annotated with colored tracks in the heatmap. Protein identifiers were mapped to ENSEMBL IDs and analyzed within the 16-layer human multiplex network described above, enabling cross-layer mechanistic inference grounded in experimentally derived biological relationships. MENTOR leverages the Random Walk with Restart (RWR) algorithm implemented in the *RWRtoolkit*[1], which quantifies topological similarity among nodes within multiplex networks. The RWR algorithm was executed in massively parallel mode on the Andes supercomputer, enabling full-network random walk exploration for each protein seed across all 16 multiplex layers simultaneously. Each protein was used as a seed node for RWR exploration, generating rank-ordered vectors of all nodes based on visitation probability. Each iteration of this high-resolution, distributed network traversal required approximately $3x10^7$ node-level probability computations, totaling roughly $10^9$ probability updates across proteins during network convergence. Vectors were truncated at the elbow of the mean rank–score curve[8], and pairwise similarities between vectors were computed using Spearman's rank correlation (ρ)**.** Dissimilarity values (1 – ρ) were hierarchically clustered using the *stats* package in R to generate a dendrogram representing topological relationships among proteins in multiplex space. Clusters (clades) were defined by cutting the dendrogram at an empirically optimized threshold (see below) to capture functionally coherent modules, with iterative subclustering applied as needed for interpretability.

**Defining clades for interpretation:** The MENTOR dendrogram represents a hierarchical map of topological and mechanistic connectivity among genes or proteins, where branch depth encodes the degree of mechanistic proximity. To ensure interpretability during AI-assisted analysis, the dendrogram was subdivided into clades of manageable size. Based on empirical benchmarking across multiplex networks, a maximum cluster size of ~15 proteins was set to preserve mechanistic coherence while avoiding functional heterogeneity. This threshold yielded 70 clades for interpretation, each capturing a locally coherent biological mechanism. Subsequent synthesis by higher-level agents integrated overlapping or complementary themes across clades to construct pathway- and system-level mechanistic narratives.

The resulting MENTOR dendrogram provided a topologically informed representation of the proteomic landscape of rehabilitation, grouping proteins into mechanistically related clades defined by shared connectivity across vascular, metabolic, inflammatory, and proteostatic pathways. These clades were annotated by integrating network topology, literature evidence, and pathway enrichment across multiplex layers to yield interpretable biological themes. Collectively, the analysis hierarchically organized the molecular response to rehabilitation at topological precision and mechanistic resolution far exceeding what can be achieved with conventional enrichment or correlation-based analyses.

**AI-assisted network interpretation:** To achieve automated, AI-assisted interpretation of these complex multiplex networks, we employed the MENTOR Interpretation Agent (MENTOR-IA)[10], a large language model (LLM)-driven, retrieval-augmented generation (RAG) interpretation system that converts network embeddings into structured mechanistic narratives. MENTOR-IA extends the MENTOR framework by automating the biological interpretation of gene or protein clades derived from random walk with restart (RWR)-based network embeddings[8,9]. The MENTOR dendrogram from our HFpEF rehabilitation proteomic targets described above was used as the primary input. Each clade, representing a topologically coherent module of mechanistically related proteins, was parsed by MENTOR-IA for systematic interpretation. The pipeline begins with retrieval augmentation for a swarm of different agents (with author, critic, and editor agents for each RAG data source), in which gene and protein identifiers from each clade are matched to multiple annotation bases (including 4,350 PubMed abstracts from the literature about these specific genes/proteins, Gene Ontology (GO) Biological Processes, GO Molecular Function, GO Cellular Compartment, KEGG, Reactome, WikiPathways, Human Protein Atlas, and NCBI mygene summaries). These materials are assembled into contextual “evidence packets” provided to the LLM (openAI GPT5) for interpretation.

Using this retrieved context, MENTOR-IA performs a multi-stage summarization process. Each clade is first processed by a set of clade interpreters (Gene-level Interpreter, Enrichment-level Interpreter, Literature Interpreter) that synthesize the retrieved evidence into concise mechanistic evidence-level summaries. An Aggregator Agent then compares and integrates these interpretations to create a clade-level mechanistic summary. Finally, a Cross-Clade Summarizer identify shared or divergent biological themes across clades, such as vascular remodeling, mitochondrial metabolism, calcium signaling, proteostasis, or immune regulation. In total, 11.8 million tokens were used as RAG input and 3.4 million tokens (≈480 pages of text) of output were generated by the LLMs. The MENTOR-IA framework was operated in a supervised, human-in-the-loop mode, where researchers reviewed and refined model outputs for accuracy, biological plausibility, and interpretability.

**Tissue-level expression of protein targets**

*GTEx*. GTEx gene expression of tissues was downloaded from human proteome atlas (https://www.proteinatlas.org/download/tsv/rna_tissue_gtex.tsv.zip, v24.0). Among the 483 proteins identified in the physical functional proteome, 469 were found to overlap with the GTEx dataset, which comprises 19,217 genes. There were 389 out of 469 genes with at least one TPM > 10 in at least one tissue. TPM values were normalized (centered and scaled) across tissues for each of those 389 genes. The top 10 tissues with the highest number of genes with z-score > 1 were visualized.

*Tabula Sapiens human cell atlas*[11]*.* Single-cell transcriptomic data were obtained from CellxGene, and 412,837 cells generated using the 10x Genomics 3′ platform were retained. Gene set activity scores were calculated using the *AddModuleScore* function in Seurat. Median activity scores across tissues or cell types were ranked and visualized.

*Human Muscle Ageing Cell Atlas (HLMA)*[12]. We analyzed 210,445 snRNA-seq cells from 15 older and 7 younger donors. Pseudobulk counts were generated at both the sample and cell-type levels. Low-abundance genes were filtered using edgeR's *filterByExpr* (min.count = 5), and differential expression was assessed using *glmLRT*. FDR correction was applied to genes shared between HLMA and the predefined gene sets. Genes with FDR < 0.05 and an absolute fold change > 1.2 were considered significant.

*Aged skeletal muscle (GSE167186)*[13]*.* Bulk RNA-seq counts and metadata for 29 older healthy, 19 younger healthy, and 24 sarcopenia subjects were downloaded from GEO.

Differential expression analysis was performed using DESeq2 for three comparisons: older healthy vs younger healthy, sarcopenia vs older healthy, and sarcopenia vs younger healthy. FDR correction was applied to genes shared between this dataset and the predefined gene sets, with significance defined as FDR < 0.1.

### SUPPLEMENTAL RESULTS

#### Agentic AI-based analysis of the rehabilitation proteomic network

The following sections describe the representative genes, signaling pathways, and biological processes underpinning these network modules, highlighting how endothelial remodeling, mitochondrial metabolism, calcium handling, sarcomeric structure and function, proteostasis, immune mechanisms, and metabolic remodeling collectively contribute to the systemic recovery phenotype observed with rehabilitation in HFpEF.

***Endothelial and Microvascular Remodeling*** (C12, C18, C24, C29, C33, C35, C37, C40, C67). This signal reflects angiogenic and antifibrotic remodeling through TGFβ–ALK1 and Notch crosstalk, supported by TGFβ1 (C35) and its receptor ACVRL1 (C35) (ALK1), which can regulate vascular morphogenesis and integrity; ACVRL1 loss causes hereditary hemorrhagic telangiectasia with disordered angiogenesis[14]. TGFβ1-mediated restraint of endothelial-to-mesenchymal transition mitigates fibrosis[15]. Up-regulation of placental growth factor (PGF) (C35) and EGFL7 (C35) in this cluster may reflect endothelial activation and capillary repair[16,17], while EGFL7 and its intronic miR-126 couple VEGF and Notch pathways to maintain vessel stability[18]. Exercise and HF studies consistently identify VEGFA as a driver of endothelial repair and microvascular growth, with skeletal-muscle VEGFA expression increasing after exercise in chronic heart failure and genetic variants influencing cardiac remodeling and outcomes[19,20]. The co-upregulation of MAPK9 with VEGFA implicates coordinated activation of MAPK/Ras signaling under stress, as in models linking MAPK9 to oxidative and inflammatory signaling cascades[21]. The inclusion of BICD1, a dynein adaptor that facilitates HIF1A nuclear translocation and hypoxia-induced VEGFA expression, situates this module within a vascular–metabolic adaptation network[22]. USP28, AMFR, and HECTD2 are involved in protein quality control and ER-associated degradation (proteostasis[23,24]). Positive VEGFA and MAPK9 estimates combined with negative BICD1 and TUT4 trends may suggest a shift from intracellular stress and RNA decay toward enhanced endothelial activation and metabolic resilience[25,26]. Although derived largely from oncology and inflammatory studies, the proteostasis genes may represent conserved stress-relief mechanisms that restore protein homeostasis during systemic rehabilitation[27,28].

***Mitochondrial Bioenergetics and Oxidative Metabolism*** (C57, C58, C59, C66, C67). FDX2 (C57) donates electrons to the Fe–S assembly machinery that supports respiratory complexes I–III and other mitochondrial enzymes. Human genetics and biochemical studies confirm that FDX2 is essential for Fe–S biogenesis, with pathogenic variants producing combined respiratory-chain deficits and myopathy[29-33]. COX5B (C57), encoding a structural subunit of cytochrome c oxidase (Complex IV), is critical to mitochondrial function, with its loss inducing mitochondrial depolarization, reactive-oxygen-species accumulation, ATP depletion, senescence, and cytokine shifts[34,35]. Together, disruption of the FDX2-COX5B axis may compromise oxidative phosphorylation[32,33], potentially coupling enhanced oxygen delivery with increased mitochondrial efficiency in rehabilitation in HFpEF. Of note, this metabolic core is coupled to calcium handling (see below) through SUMO1 (C65), which stabilizes SERCA2a thereby potentially linking mitochondrial energetics to relaxation and excitation–contraction mechanisms relevant to SPPB and 6MW.

***Calcium Handling and Excitation–Contraction Coupling***. The calcium-handling module bridges mitochondrial and contractile sectors, featuring a link from SUMO1 (C57), to CACNB1 (C65), CRACR2B (C65), and TRPC4AP (C65). SUMO1 SUMOylates SERCA2a, stabilizing the pump and enhancing sarcoplasmic $Ca^{2+}$ reuptake; SUMO1 gene transfer restores cardiac contractility in failing hearts[36,37]. CACNB1 promotes assembly of L-type $Ca^{2+}$ channels, and TRPC4AP organizes transient receptor potential channel complexes involved in endothelial $Ca^{2+}$ entry[38,39]. This mechanistic theme likely illustrates the functional coupling between energetic and contractile systems, coordinating mitochondrial activity and calcium cycling to enhance diastolic relaxation and excitation–contraction synchrony in trained participants.

**Sarcomeric Structure and Mechanotransduction.** The sarcomeric and mechanotransductive clades include TTN (C20), OBSCN (C68), and ANKRD2 (C20), reflecting remodeling of contractile stiffness and structural elasticity. TTN isoform shifts alter passive tension and diastolic compliance[40]. OBSCN maintains myofibrillar alignment and $Ca^{2+}$-related organization, and its disruption leads to disordered sarcomeric assembly[41-44]. ANKRD2 acts as a stretch-responsive regulator, potentially linking mechanical load to transcriptional adaptation[45,46]. The adjacency of this sector to the calcium branch reflects a mechanical–electromechanical continuum driving adaptive contractile remodeling with rehabilitation.

**Proteostasis and ER–Mitochondrial Crosstalk.** At the interface between the mitochondrial and sarcomeric quadrants, the proteostasis and ER-mitochondrial crosstalk clade encompasses EDEM2 (C27) and CKAP4 (C27), two genes coordinating quality control and organelle communication. EDEM2 initiates ER-associated degradation (ERAD) via glycoprotein trimming, facilitating clearance of misfolded proteins[47]. CKAP4 tethers the ER to mitochondria and mediates DKK1/3–PI3K–Akt signaling that regulates metabolism and endothelial integrity[48-50]. This cluster indicates that proteostatic adaptation may act to integrate sustaining energy metabolism and $Ca^{2+}$ homeostasis during heightened metabolic demand in rehabilitation.

**Extracellular Matrix Remodeling and Fibrosis.** The extracellular matrix (ECM) remodeling and fibrosis module spans multiple clades (C3, C14, C17, C29, C37, C45), reflecting coordinated regulation of matrix composition, turnover, and mechanical signaling across distinct but interconnected ECM subprograms. Structural ECM components and elasticity regulators, including COL6A6 (C3) and ELN (C45), contribute to baseline tissue stiffness and compliance, influencing passive myocardial and vascular mechanics through collagen VI organization and elastic fiber integrity[51,52]. Protease restraint mechanisms captured in C14, most prominently RECK, limit excessive matrix metalloproteinase activity during remodeling, thereby preserving ECM integrity and promoting controlled matrix turnover rather than pathological fibrosis[53]. Regulation of TGFβ signaling emerges through ECM-associated binding and elastic fiber networks in C17, including LTBP3, which modulates sequestration and activation of latent TGFβ complexes and links matrix organization to fibroblast activation and fibrotic signaling[54]. Matricellular signaling proteins in C29, such as CCN1 (CYR61), further couple ECM structure to integrin-dependent mechanotransduction, fibroblast activation, and vascular–matrix crosstalk in response to mechanical stress, integrating extracellular stiffness with cellular signaling responses[55]. Additional ECM remodeling signals in C37, including LOXL2 and the hypoxia-responsive transcription factor EPAS3 (HIF-2α), link collagen and elastin cross-linking with hypoxia-adaptive gene regulation, providing context-dependent control of matrix stiffness and vascular remodeling under metabolic or inflammatory stress[56,57]. Collectively, these clades define an adaptive ECM program in which matrix structure, signaling, and turnover are dynamically regulated during rehabilitation, promoting matrix plasticity that balances structural support with restored compliance to enable improved vascular function and myocardial mechanics.

**Innate Immune and Myeloid Activation/Resolution.** CSF3R (C15), the receptor for G-CSF, controls granulopoiesis and neutrophil activation[58]. NCF2 (p67-phox) (C15), a NADPH-oxidase subunit, is required for the respiratory burst; its dysfunction reduces reactive oxygen species generation and inflammatory activation[59]. LAIR-2 (C15) acts as a soluble decoy for LAIR-1, binding collagen and mitigating inhibitory signaling[60]. CD300e (IREM-2) (C15) drives monocyte and dendritic-cell activation[61], and VNN2 (GPI-80) (C15) supports $\beta_2$-integrin-dependent adhesion and transendothelial migration[62]. The topological co-localization of these immune genes suggests a coordinated down-regulation of pro-inflammatory and oxidative pathways, consistent with exercise-induced immune normalization in HFpEF[63].

**Anti-Atrophy and Aging-Linked Signaling.** The EDA2R (XEDAR) (C1) cluster represents the anti-atrophy and aging-linked signaling domain. EDA2R activates non-canonical NF-κB pathways that promote muscle catabolism; suppression preserves mass in aging and cachexia models[64]. Its down-regulation post-intervention aligns with reduced sarcopenic signaling and restoration of anabolic balance, corresponding to functional strength gains observed in rehabilitation responders.

**Endocrine and Metabolic Regulation.** This theme includes adipocyte remodeling and lipid-droplet dynamics with CLSTN3β (C53) as a defining node. This adipocyte-specific isoform localizes to ER–lipid-droplet contact sites, restricting droplet fusion and fostering multilocular, oxidative adipocytes[65]. This combined with mitochondrial mechanisms likely indicates integration of lipid mobilization and oxidative metabolism, providing a substrate foundation for endurance and metabolic resilience with rehabilitation.

**Supplemental Table 8. Proteomic relations to physical function:** Table of proteins selected by LASSO models for SPPB and 6-minute walk distance. Top effect proteins were selected for discussion.

| Protein | Relation to physical function | Function |
|---|---|---|
| ACAD9 | + | Key factor in mitochondrial metabolism[66]. Deficits in ACAD9 are associated with cardiomyopathy, weakness and exercise intolerance[67]. |
| AGR3 | + | Involved in epithelial homeostasis and secretion. No established roles with frailty or CVD. |
| ASAH2 | + | Key enzyme in metabolism of ceramide, which in turn affects cell division, apoptosis and senescence[68]. Dysfunction in ceramide or sphingolipid metabolism leads to mitochondrial dysfunction, insulin resistance and inflammation[69]. |
| C4orf17 | - | Transmembrane protein, poorly understood mechanism. No established roles with frailty or CVD. |
| CALB2 | - | Regulates lipid metabolism. May be causally implicated in abdominal aortic aneurysm development[70]. |
| CCDC73 | - | Involved in ciliary movement[71]. No established role in frailty or CVD. |
| CD300E | - | Transmembrane protein expressed on myeloid cells, involved in both adaptive and innate immune responses[72]. No established role in frailty or CVD. |
| CDHR5 | + | Expressed in epithelial cells of gut and kidney where its role is microvilli organization and epithelial polarity. While not directly implicated in CVD, cadherins in general are involved in proliferation and survival of cardiomyocytes, vascular smooth muscle cells and endothelial cells[73]. |
| CPA2 | + | Involved in post-translational modification of proteins. May also be involved in lipid metabolism[74]. |
| DCXR | + | Involved in carbohydrate metabolism and detoxification of a-dicarbonyl compounds[75]. Accumulation of these compounds is linked to vascular endothelial dysfunction, atherogenesis and tissue senescence[76]. |
| EDN1 | - | Potent vasoconstrictor produced by vascular endothelial cells. Promotes vascular dysfunction, inflammation and cellular senescence; key factors in CVD and frailty[77,78]. |
| ERBB2 | + | Tyrosine kinase receptor essential for cardiac homeostasis. Disruption of this signaling pathway may lead to cardiomyopathy[79]. Negatively genetically associated with accelerated cardiac aging[80]. |
| FDX2 | - | Central regulator of lipid metabolism. Deficiencies cause lipid accumulation such as in hepatic steatosis[81]. |
| FGFBP2 | + | Modulates fibroblast growth factor signaling; preferentially secreted by cytotoxic lymphocytes. |

| | | |
|---|---|---|
| FLT3LG | + | Regulates development of dendritic cells. Implicated in vascular calcification[82]. |
| GH1 | - | Central mediator of growth. Regulates skeletal muscle maintenance. As GH1 levels decrease with age, body composition changes to include more fat mass and less lean muscle[83]. Complex relationship with CVD as both excess and deficiency cause adverse effects[84]. |
| GOLGA4 | + | Facilitates interactions between the Golgi apparatus and microtubules. No established role in frailty or CVD. |
| IL10RA | + | Involved in anti-inflammatory signaling. Deficiencies in IL10/IL10RA pathways results in frailty, cardiac fibrosis and hypertrophy, and vascular remodeling[85,86]. |
| IL15 | - | Inflammatory cytokine. Higher levels associated with atherosclerotic disease[87]. Alterations seen in pre-frail and frail individuals[88]. |
| IL6 | - | Inflammatory cytokine. Elevated levels of IL6 are related to greater risk of frailty and cardiovascular disease (including heart failure)[89]. Emerging evidence on therapeutic inhibition may be beneficial in CVD prevention[90]. |
| LIFR | - | Inflammatory cytokine. Genetic analysis implicated LIFR in the pathogenesis of frailty[91]. It is also involved in atherosclerosis[92]. |
| LRRN1 | + | Transmembrane protein involved in neurogenesis and synaptic activity. No established role in frailty or CVD. |
| MKI67 | - | Nuclear scaffolding protein which is only active during cell proliferation. No established role in frailty or CVD. |
| MSTN | + | Negatively regulates muscle growth; contributes to muscle wasting and cachexia[93]. Increased in heart failure and may be related to cardiac remodeling[94]. Implicated in vascular dysfunction[95]. |
| NEFL | – | Structural protein on the neuronal axoskeleton. Higher levels are associated with frailty and CVD[96,97]. |
| N5T5C1A | - | Cytosolic nucleotidase. No established role in frailty or CVD. |
| PLIN1 | - | Regulates lipid accumulation and plaque stability[98]. In cardiac tissue, modulates responses to oxidative stress and may cause hypertrophy[99,100]. |
| PXDC1 | + | May be involved in epigenetic regulation[101]. Identified in a GWAS of frailty, but its role is unclear[102]. |
| SDHA | - | Central to mitochondrial metabolism. Decreased activity seen in heart failure models[103]. Not directly implicated in frailty, but abnormal mitochondrial energetics are a common driver of frailty[85]. |
| SMPDL3A | + | Regulates inflammatory signaling and involved in atherosclerosis[104]. |
| SYTL1 | - | Central to exocytosis and linked to chemotaxis of neutrophils. No established role in frailty or CVD. |
| TFDP3 | - | Involved in cell cycle regulation. No established role in frailty or CVD. |

| | | |
|---|---|---|
| TPX2 | - | Microtubule associated protein involved in cell cycle regulation and DNA repair. No established role in frailty or CVD. |

**Supplemental Figure 1. Analytical design.**

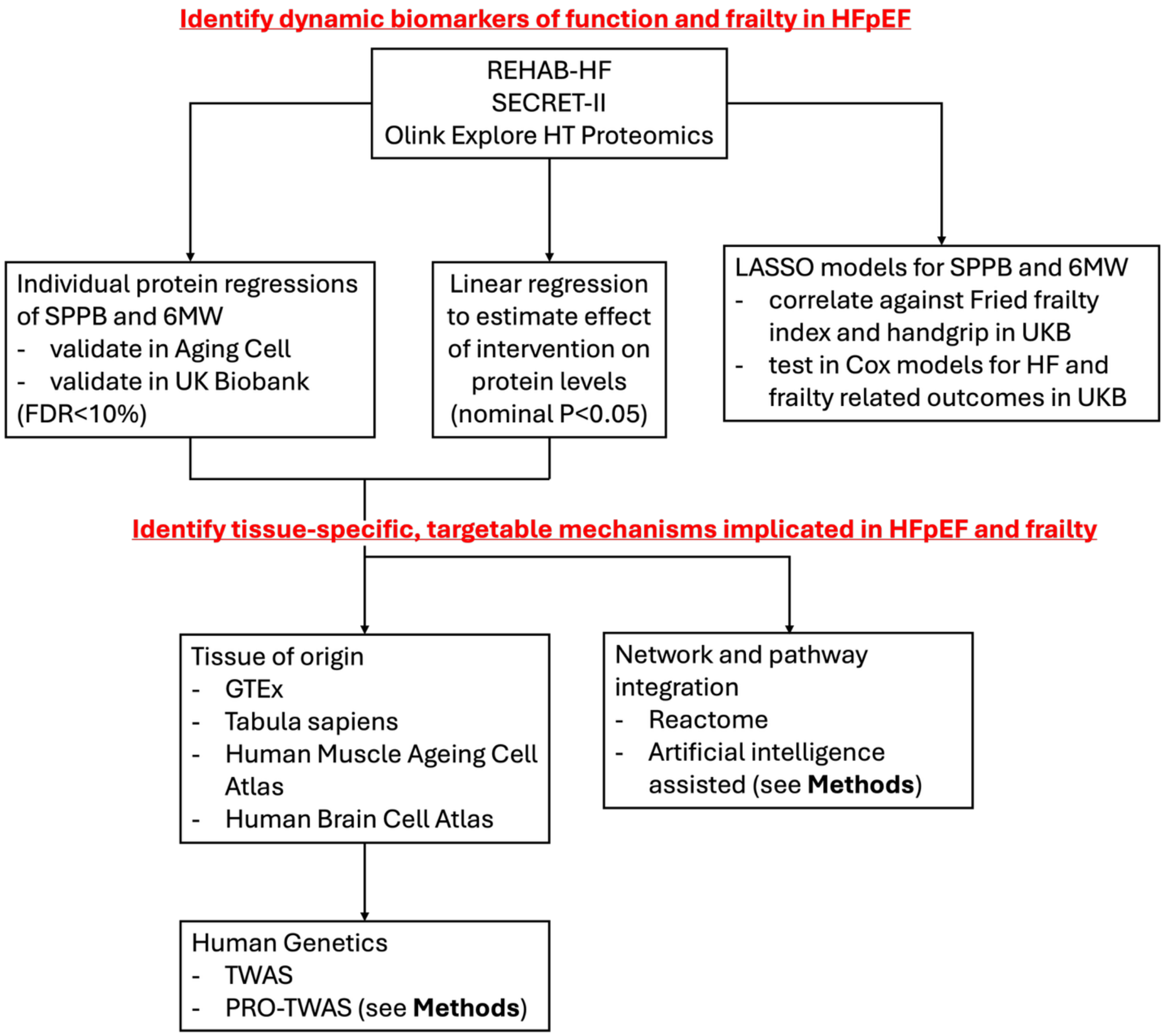

**Supplemental Figure 2. Distribution of short physical performance battery (SPPB) and 6-minute walk distance (6MW) in REHAB-HF and SECRET-II.** (A) Density plot of SPPB stratified by trial. (B) Density plot of 6MW stratified by trial.

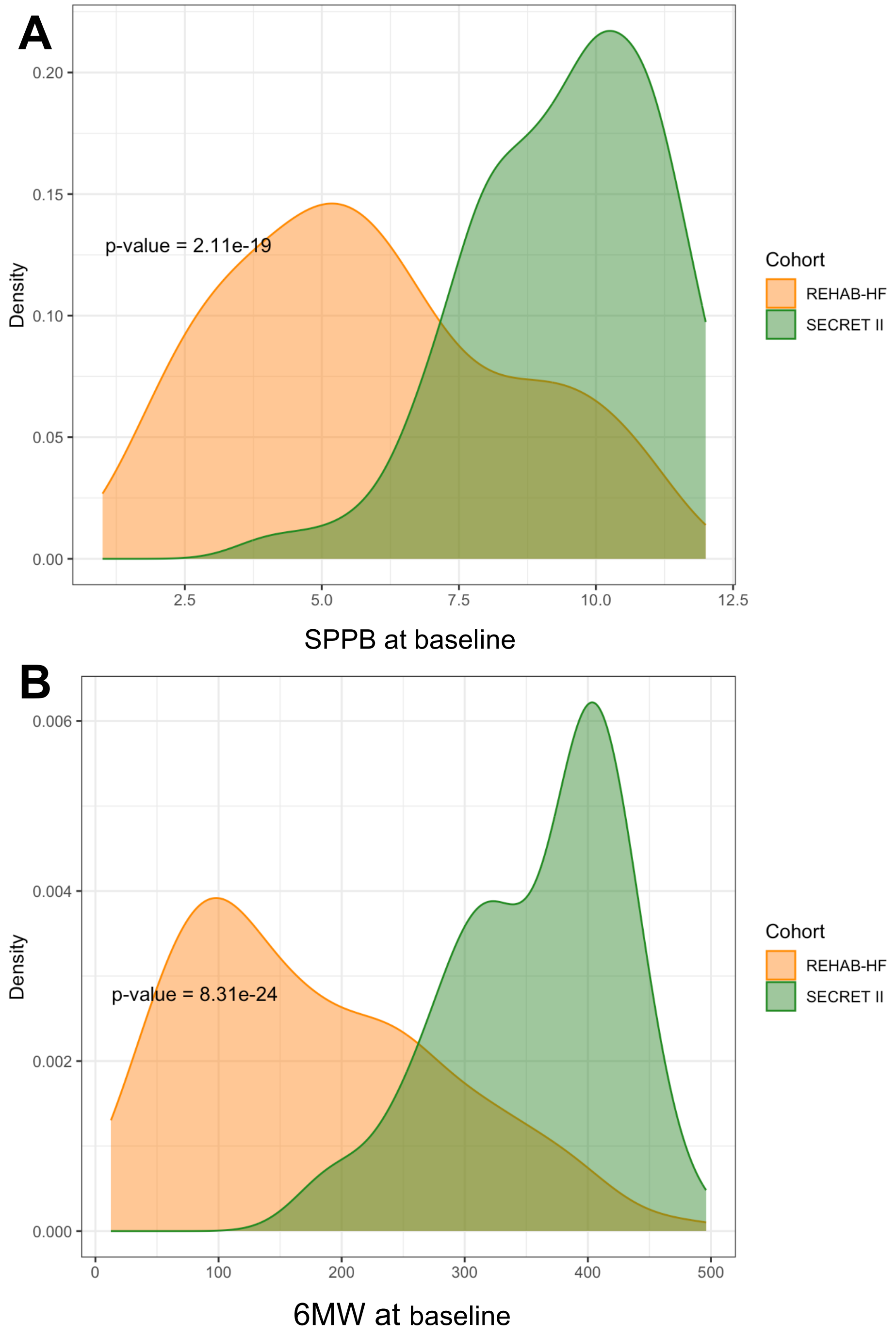

**Supplementary Figure 3. Over-Representation analysis.** For proteins associated with SPPB, 6MW in REHAB-HF and SECRET-II at FDR<10% and proteins with changed expression after intervention in REHAB-HF or SECRET-II at a nominal P<0.05, we show the top 50 enriched Reactome pathways using "genome protein-coding" as reference set.

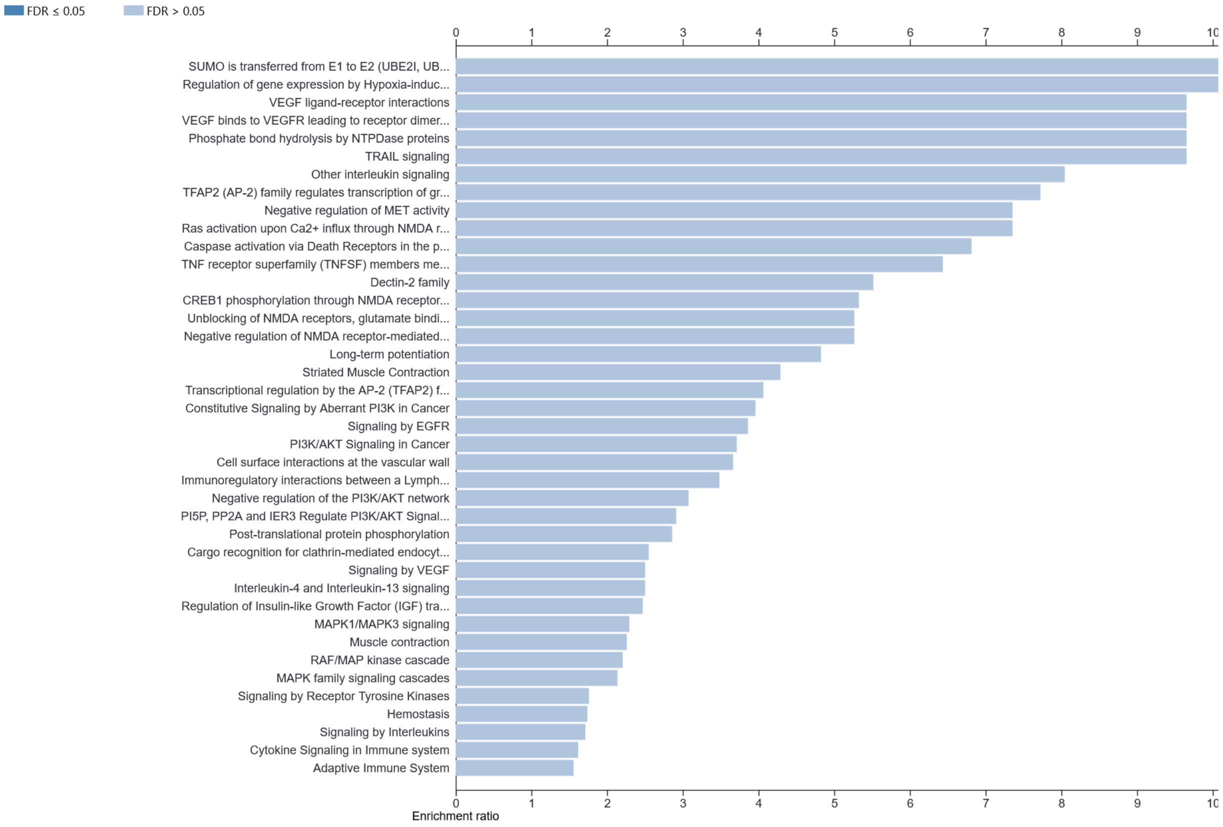

**Supplemental Figure 4. Tissue and cell-specific localization of the physical function related proteome.** (A) Gene activity scores of 6MW/SPPB-associated proteins across tissues in the Tabula Sapiens dataset. (B) Gene activity scores of intervention-associated proteins across tissues in the Tabula Sapiens dataset. (C) Gene activity scores of 6MW/SPPB-associated proteins in the top 25 cell types with the highest median gene activity in the Tabula Sapiens dataset. (D) Gene activity scores of intervention-associated proteins in the top 25 cell types with the highest median gene activity in the Tabula Sapiens dataset.

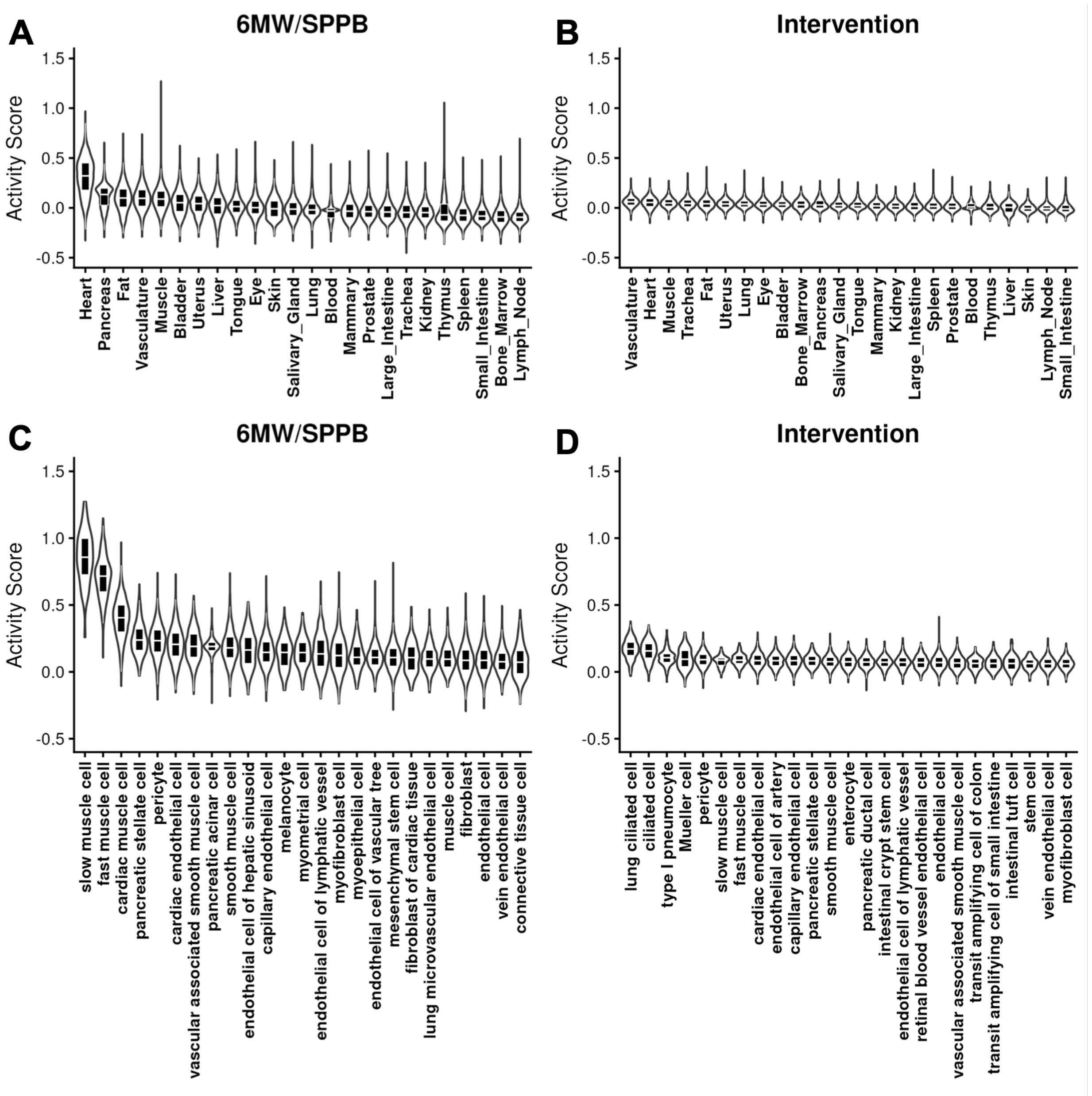
A
6MW/SPPB
B
Intervention
C
6MW/SPPB
D
Intervention
Activity Score
Heart
Pancreas
Fat
Vasculature
Muscle
Bladder
Uterus
Liver
Tongue
Eye
Skin
Salivary_Gland
Lung
Blood
Mammary
Prostate
Large_Intestine
Trachea
Kidney
Thymus
Spleen
Small_Intestine
Bone_Marrow
Lymph_Node
slow muscle cell
fast muscle cell
cardiac muscle cell
pancreatic stellate cell
pericyte
cardiac endothelial cell
vascular associated smooth muscle cell
pancreatic acinar cell
smooth muscle cell
endothelial cell of hepatic sinusoid
capillary endothelial cell
melanocyte
myometrial cell
endothelial cell of lymphatic vessel
myofibroblast cell
myoepithelial cell
endothelial cell of vascular tree
mesenchymal stem cell
fibroblast of cardiac tissue
lung microvascular endothelial cell
muscle cell
fibroblast
endothelial cell
vein endothelial cell
connective tissue cell
lung ciliated cell
ciliated cell
type I pneumocyte
Mueller cell
enterocyte
pancreatic ductal cell
intestinal crypt stem cell
retinal blood vessel endothelial cell
transit amplifying cell of colon
transit amplifying cell of small intestine
intestinal tuft cell
stem cell
myofibroblast cell

**Supplemental Figure 5. Differential expression status of 6MW/SPPB-associated proteins in the aging human muscle atlas.** (A) Numbers of differentially expressed and non–differentially expressed genes. (B) Proportion of differentially expressed genes.

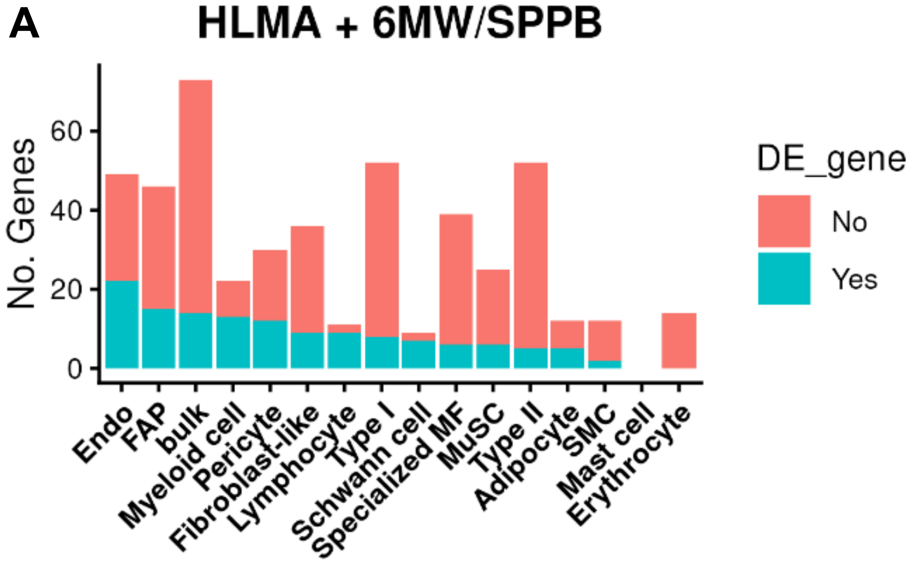


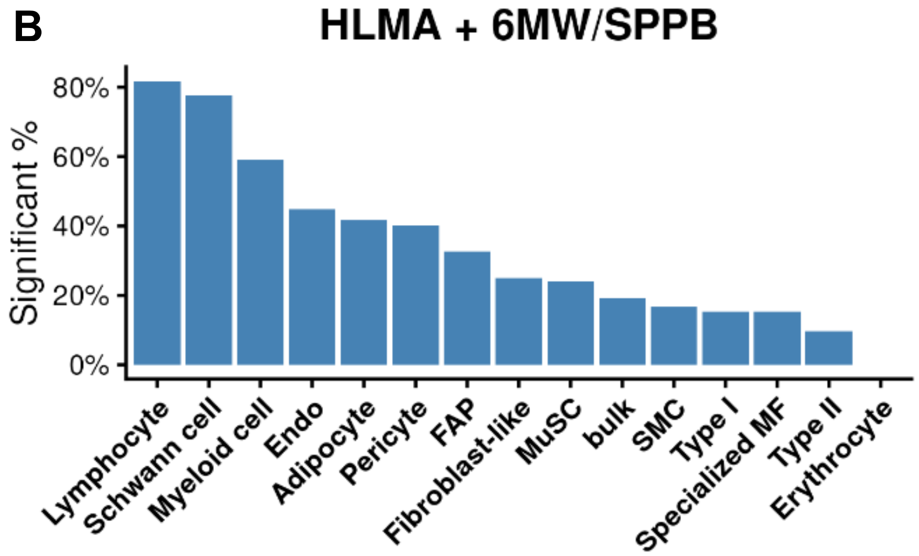

**Supplemental Figure 6. Cross-tissue significance of the physical function proteome by TWAS approaches.** The columns show the –log10 transformed p-values from the rehabilitation proteomic analysis and the TWAS analysis in heart, brain, and muscle.

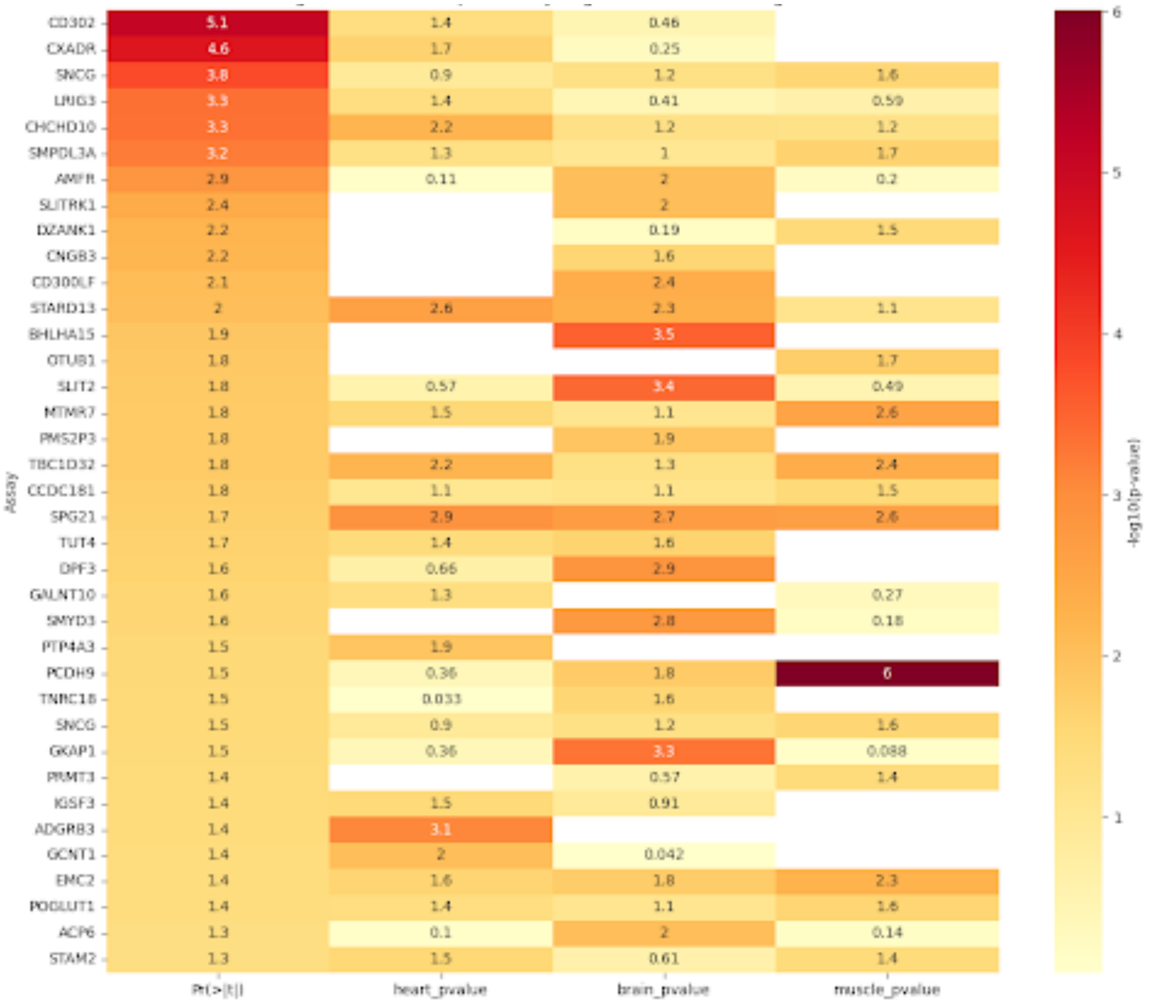

**Supplemental Figure 7. Correlation between protein scores of physical function. (A, B)** Spearman correlation of the SPPB and 6MW protein scores at the follow-up exams in REHAB-HF and SECRET-II trials. **(C, D)** Spearman correlations between proteins scores and peak oxygen consumption (VO2) in SECRET-II. **(E, F)** Spearman correlation between protein scores and handgrip in UK Biobank. Spearman correlation. SPPB = Short Physical Performance Battery; 6MW = 6-minute walk.

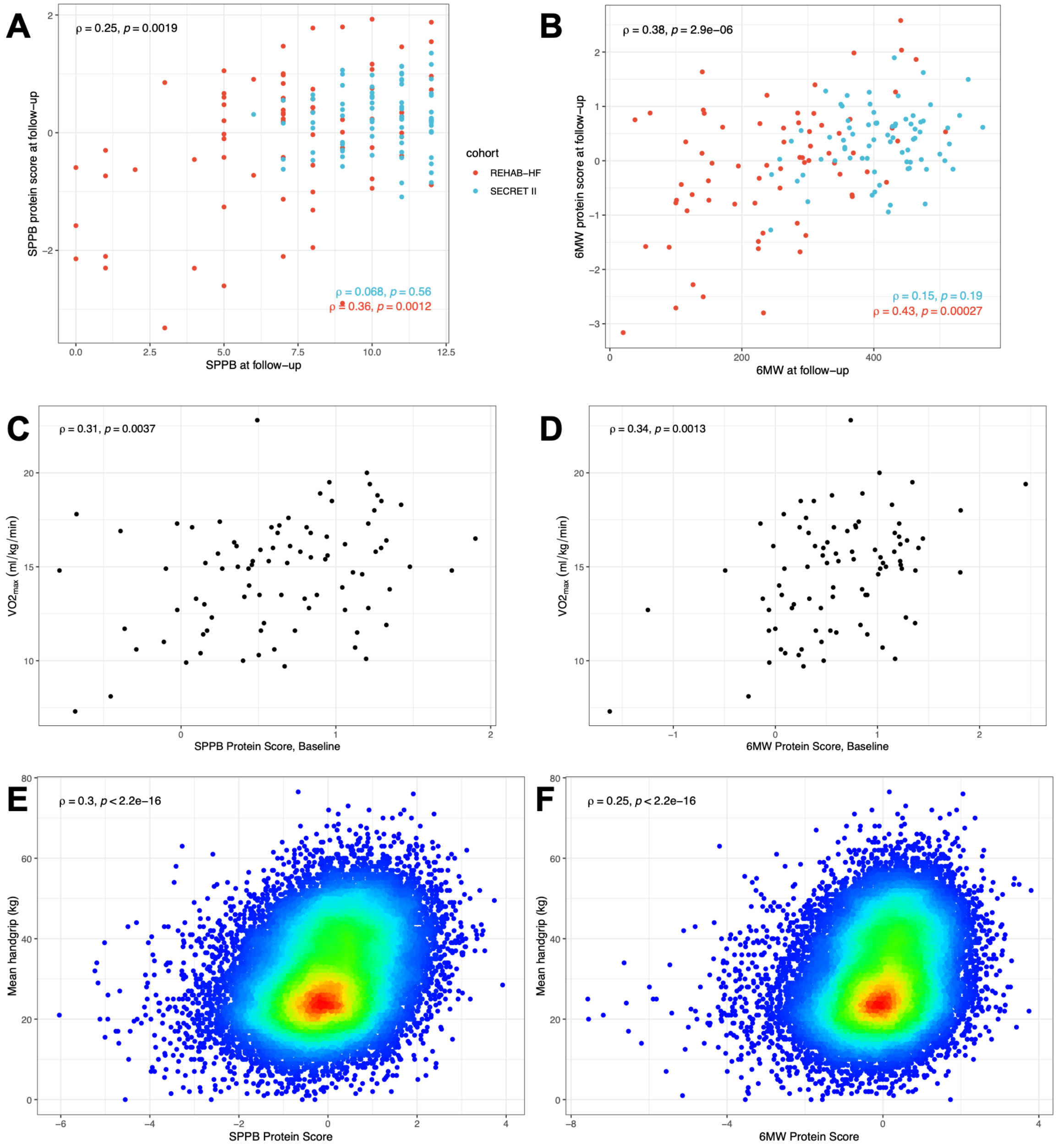

**Supplemental Figure 8. Model fit of recalibrated protein scores for use in UK Biobank.** Given the reduced proteomic coverage in UK Biobank (≈3,000) compared to our discovery cohort (≈5,000) we performed LASSO regressions on the proteins common to both cohorts for the outcome of the original protein scores.

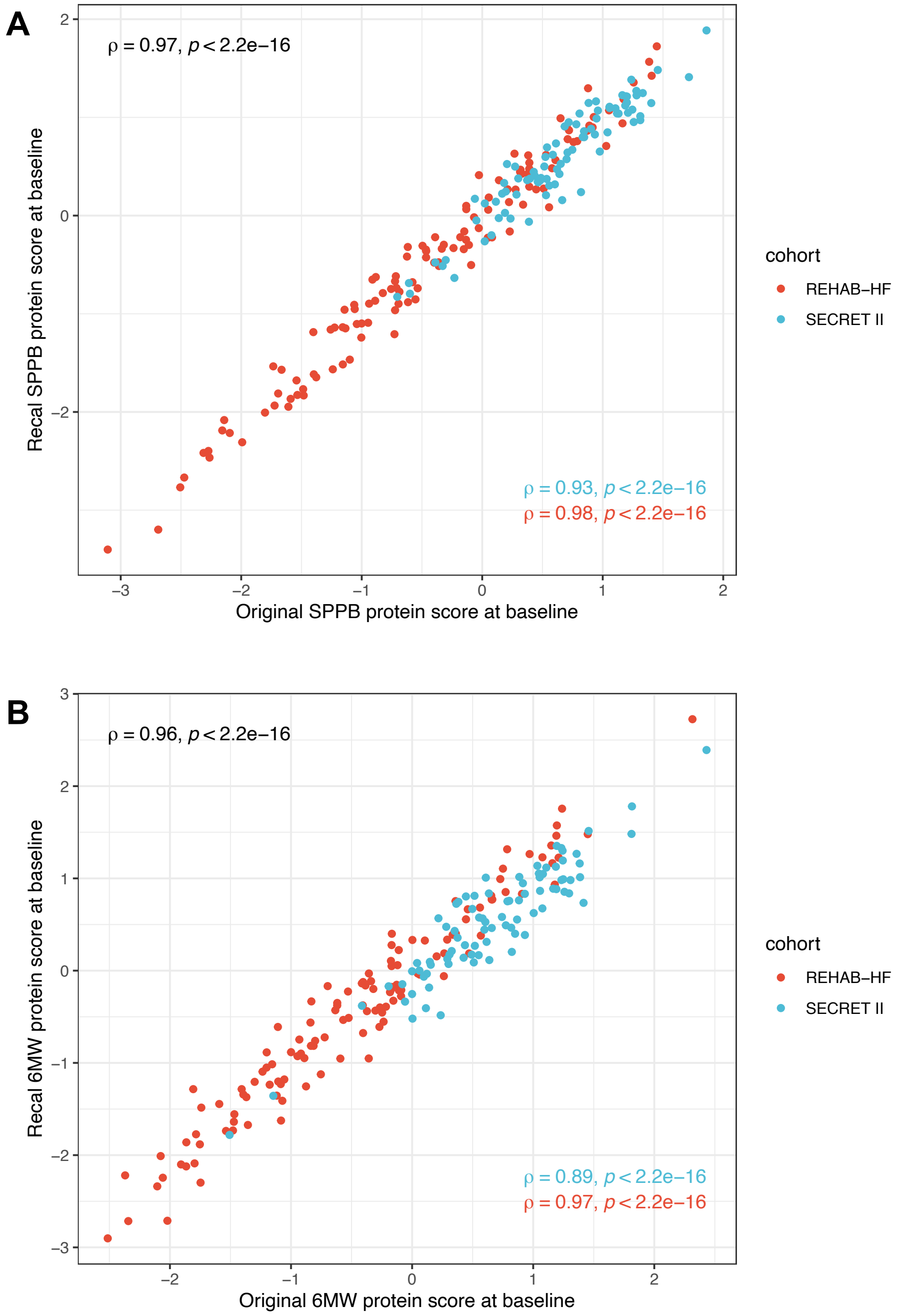
